# Carbonaceous Components Underlying Wildfire Smoke PM_2.5_-Associated Alzheimer’s Disease and Related Dementias in Older US Adults

**DOI:** 10.64898/2026.10.08.26364863

**Authors:** Zhihao Jin, Ke Xu, Meng Qi, Qiao Zhu, Binyu Yang, Gonzalo A. Ferrada, Joshua S. Fu, Noah Scovronick, Howard H. Chang, Alvaro Alonso, Kyle Steenland, Yang Liu

## Abstract

Wildfire smoke exposure is increasing across the U.S., but the long-term neurological effects of smoke-specific fine particulate matter (PM_2.5_) and its carbonaceous components remain uncertain. Using Medicare data from 2002 through 2019, we examined incident Alzheimer’s disease (AD) and Alzheimer’s disease and related dementias (ADRD) among 39,678,630 fee-for-service Medicare beneficiaries followed from 2005 through 2019. We identified 3,501,414 incident AD cases over 258.5 million person-years and 8,377,416 incident ADRD cases over 248.6 million person-years. We linked the cohort with annual, high-resolution estimates of smoke and non-smoke PM_2.5_ mass and carbonaceous components (elemental and organic carbon) and fitted stratified Cox proportional hazards models. Per 1 µg/m^3^ increase, smoke PM_2.5_ was associated with incident AD (hazard ratio [HR], 1.015, 95% confidence interval [CI], 1.005-1.026) and ADRD (1.029 [1.024-1.035]). Smoke carbonaceous fraction was also associated with AD (1.063 [1.042-1.085]) and ADRD (1.090 [1.077-1.103]). Within the ADRD endpoint, smoke-related associations exceeded their corresponding non-smoke associations on a per-unit-mass basis. Per interquartile range, associations were smaller for smoke PM_2.5_ (AD, 1.005 [1.002-1.008]; ADRD, 1.009 [1.008-1.011]) than for non-smoke PM_2.5_ (AD, 1.086 [1.079-1.094]; ADRD, 1.048 [1.043-1.053]). Associations varied across geographic, racial, socioeconomic, and clinical subgroups. Long-term wildfire smoke exposure, particularly its carbonaceous fraction, was associated with higher incidence of AD and ADRD among older U.S. adults. The associations correspond to 2,284 and 11,142 incident cases per year of AD and ADRD attributable to smoke PM_2.5_ among older Medicare beneficiaries, with the carbonaceous fraction accounting for most of this burden. These findings support the use of source-resolved and composition-specific smoke metrics in dementia research and wildfire-smoke health-impact assessments.

## Introduction

Wildland fire activity across the U.S. has intensified substantially in recent decades, with increases in burned area, fire intensity, and fire-season length.^1,2^ Anthropogenic climate change has amplified the meteorological and ecological conditions that promote fire spread and persistence, including elevated temperatures, reduced fuel moisture, prolonged drought, and altered precipitation patterns.^3,4^ These shifts have made air pollution from wildfire smoke a major pathway through which climate change affects population health in the U.S.^5^ Wildfire smoke is a chemically complex combustion pollution mixture composed of particulate and gaseous pollutants, among which fine particulate matter (PM_2.5_) is the principal pollutant of concern for public health.^6^ Growing toxicological evidence indicates that inhaled smoke-derived particulate matter may affect the central nervous system through multiple pathways, including direct translocation through the olfactory neural system, disruption of the blood-brain barrier through systemic and neurovascular inflammation, and activation of oxidative and inflammatory cascades implicated in neurodegenerative processes.^7,8^

Alzheimer’s disease and related dementias (ADRD) represent a defining public health challenge among aging populations. In 2024, approximately 6.9 million Americans aged 65 years and older were living with Alzheimer’s disease (AD), a number projected to increase to 13.8 million by 2060, with annual health care costs exceeding $300 billion.^9^ Although established risk factors, including advanced age, APOE-ε4 genotype, hypertension, diabetes, and other cardiometabolic and behavioral conditions, explain part of the population burden, a substantial fraction of ADRD risk remains unexplained, underscoring the importance of modifiable environmental contributors.^10^ Long-term exposure to ambient PM_2.5_ has emerged as a key contributor. In a national Medicare cohort of 12.2 million older adults followed from 2000 through 2018, each interquartile range (IQR) increase in 5-year average PM_2.5_ (3.2 µg/m^3^) was associated with a 6.0% increase in incident dementia (hazard ratio [HR]: 1.060; 95% confidence interval [CI]: 1.054-1.066) and a 7.8% increase in incident AD (1.078 [1.070-1.086]).^11^ Parallel evidence has linked long-term PM_2.5_ exposure to first hospital admissions for neurological disorders and to greater neuropathologic and clinical severity among individuals with AD or dementia.^12–14^ However, most epidemiologic evidence has focused on total ambient PM_2.5_, which may obscure source-specific toxicity. Evidence specific to smoke-derived PM_2.5_ is more limited. In the Health and Retirement Study, Zhang et al. reported that source-apportioned wildfire PM_2.5_ was associated with incident dementia after adjustment for PM_2.5_ from other sources and co-pollutants (HR per IQR, 1.05; 95% CI, 1.02-1.08).^15^ A recent study by Do et al. among Medicare beneficiaries with prevalent ADRD reported largely null associations between short-term wildfire smoke PM_2.5_ and acute hospitalization, but this study did not address long-term exposure and ADRD incidence.^16^ More recently, Elser et al. evaluated 3-year average wildfire PM_2.5_ in relation to incident dementia and found no significant association.^17^ Altogether, evidence remains limited, and no national cohort study has examined long-term exposure to smoke-derived PM_2.5_ as a risk factor for incident AD and ADRD.

A further question is which specific chemical components of smoke PM_2.5_ are responsible for the smoke-dementia association. Although the constituents of PM_2.5_ (e.g., carbonaceous matter, secondary inorganic aerosols such as nitrates and sulfates, metals, and mineral particles) have each been associated with adverse health impacts, the relative magnitudes of these associations can vary across source mixtures, populations, and regions.^18,19^ Compared with non-smoke PM_2.5_, wildfire smoke is enriched in carbonaceous material generated through the combustion and incomplete burning of organic materials.^20,21^ These carbonaceous constituents include organic carbon (OC), which is derived largely from partially burned biomass and secondary organic aerosol formation, and elemental carbon (EC), which is formed primarily during incomplete combustion.^21^ Component-specific epidemiologic evidence supports the relevance of PM_2.5_ chemical composition for ADRD. Using weighted quantile sum regression and quantile g-computation, Zhang et al. identified long-term exposure to sulfate, OC, and copper as the PM_2.5_ components most strongly associated with incident AD and dementia.^22^ This finding suggests that the carbonaceous fraction of ambient PM_2.5_ is implicated in dementia pathogenesis, but the analysis did not distinguish the sources of these components. OC and EC also originate from non-smoke sources (e.g., fossil-fuel combustion, traffic, and industry) and coexist with different non-carbonaceous components (e.g., sulfate, nitrate, metals).^23,24^ These source-specific differences in chemical mixtures suggest that smoke-derived and non-smoke OC and EC may not be toxicologically equivalent per unit mass. Consistent with this, we recently found in a national Medicare cohort that the carbonaceous fraction of smoke PM_2.5_ accounted for most of the smoke-associated mortality burden.^25^ However, whether long-term exposure to wildfire smoke OC and EC increases the risk of AD or ADRD in the general older adult population remains unaddressed in a national, source-resolved, chemically resolved cohort framework. This gap largely reflects the limited availability of long-term, high-resolution exposure estimates that distinguish smoke from non-smoke sources and characterize the chemical composition of smoke PM_2.5_.

To address these gaps, we linked a national cohort of U.S. Medicare beneficiaries aged 65 years and older from 2002 through 2019 with high-resolution estimates of smoke-specific PM_2.5_ mass, OC, and EC. The primary objectives of the study were to quantify associations of 3-year moving-average smoke PM_2.5_ mass and smoke carbonaceous components (carbon, OC+EC) with incident AD and incident ADRD using stratified Cox proportional-hazards models, with mutual adjustment for non-smoke counterparts. We also aimed to assess effect modification by sex, race, Medicaid eligibility, entry age, geographic region, and three baseline comorbidities, and estimate the AD and ADRD burden attributable to smoke PM_2.5_ and smoke carbon. This study provides source-resolved epidemiological evidence to inform climate adaptation, air quality regulation, wildfire smoke mitigation, and dementia prevention in an aging U.S. population.

## Methods

### Study design and data sources

We conducted a nationwide open-cohort study by linking two data sources from the Centers for Medicare and Medicaid Services (CMS). The Medicare denominator file provided annual individual-level information on demographic characteristics, enrollment status, residential location, and vital status. The Chronic Conditions Warehouse (CCW) provided predefined claims-based chronic condition indicators and the earliest recorded diagnosis date for predefined conditions among fee-for-service beneficiaries. This study was conducted under a data use agreement with the Centers for Medicare & Medicaid Services. Informed consent was waived because the study involved a secondary analysis of existing administrative data.

### Study population

Eligible participants were Medicare beneficiaries aged 65 to 100 years who resided in the contiguous U.S. (CONUS) and were continuously enrolled in fee-for-service Medicare with both Part A and Part B coverage. These restrictions were applied because the CCW chronic conditions algorithm identifies qualifying diagnoses from fee-for-service (FFS) claims submitted through Part A and Part B claim streams. We constructed two cohorts for incident AD and incident ADRD from 2002 through 2019. For each outcome, beneficiaries were required to complete a 3-year washout period after initial enrollment, during which no qualifying diagnosis could be recorded. The purpose of this washout period was to exclude prevalent disease from the analysis, in order to focus on incident disease. Analytic follow-up began in the calendar year after completion of the washout period. Consequently, the minimum age at cohort entry was 68 years. The washout period was excluded from analytic person-time, because individuals were not yet at risk of an incident outcome during this period, thereby avoiding immortal-time bias.^26^ Follow-up began no earlier than January 1, 2005, to complete the washout period and also to ensure complete 3-year exposure histories. Participants were followed until the outcome of interest, death, disenrollment, or December 31, 2019.

### Outcomes: incident AD and ADRD

Two primary outcomes were examined: incident AD and incident ADRD. We identified AD and ADRD using CCW predefined indicators, which are based on claims-based algorithms that incorporate diagnoses from multiple Medicare claims sources. Medicare claims algorithms for dementia have shown reasonable validity in prior studies, although diagnostic sensitivity varies by dementia subtype and disease severity.^27,28^ Detailed International Classification of Diseases (ICD) codes used to define both outcomes are provided in Table S1.

We constructed two outcome-specific inception cohorts using independent 3-year washout periods, allowing beneficiaries to contribute person-time to both cohorts if they were free of the corresponding outcome at cohort entry. For the ADRD cohort, incident ADRD was defined as the first recorded qualifying diagnosis for either AD or ADRD. For the AD cohort, incident AD was defined as either the first recorded qualifying AD diagnosis among beneficiaries with no prior ADRD diagnosis, or the first recorded ADRD diagnosis among beneficiaries who subsequently received an AD diagnosis. In the latter case, the initial ADRD diagnosis was treated as the incident AD date because it was considered likely to represent an early nonspecific diagnosis of AD.

### Exposure assessment

Daily 1-km concentrations of smoke-specific and non-smoke PM_2.5_ mass, OC, and EC from 2002 through 2019 were obtained from the prediction framework developed by Jin et al.^2^ This residual-adjusted super learner integrated satellite aerosol optical depth retrievals, chemical transport model outputs from Community Multiscale Air Quality Model (CMAQ), ground-based monitor data (3,081 and 410 monitoring stations for PM_2.5_ and OC/EC, respectively), and auxiliary spatial and spatiotemporal predictors. Smoke-attributable concentrations were estimated as the difference between smoke-impacted and non-smoke predictions. Ten-fold cross-validated annual R^2^ values were 0.979 to 0.980 for OC, 0.980 to 0.985 for EC, and 0.901 to 0.939 for PM_2.5_ mass. Annual ZIP code-level concentrations were derived by population-weighted spatial averaging of daily 1-km predictions, using LandScan gridded population data.^29^ We defined PM_2.5_ carbonaceous components (carbon) as the sum of OC and EC, which was selected as the primary carbonaceous exposure because separate OC and EC estimates may be sensitive to collinearity (Table S2) and shared measurement uncertainty. The combined metric was not intended to imply that OC and EC are biologically equivalent. Non-smoke counterparts were defined analogously. For each beneficiary-year, we defined chronic exposure as the 3-year moving average across the three preceding calendar years. For example, exposure assigned to follow-up year 2011 was calculated as the mean of annual concentrations from 2008 through 2010. This same averaging window was applied to all primary exposure metrics, including smoke PM_2.5_, smoke carbon, and non-smoke counterparts.

### Covariates

For each beneficiary-year, we obtained individual-level demographic and enrollment information, together with residential ZIP code. Models accounted for these characteristics by stratifying the baseline hazard by sex, race, 5-year age-at-entry group, and Medicaid dual-eligibility status, which served as an individual-level indicator of socioeconomic status. Attained age was included as an annually updated time-varying covariate. Annually updated ZIP code-level covariates were obtained from the U.S. Census Bureau and the American Community Survey and included five neighborhood characteristics: proportion of Black residents, proportion of residents living in rental housing, proportion of residents living below the federal poverty line, proportion of residents aged 65 years or older, and residential population. We further adjusted for ZIP code-level 3-year moving averages of temperature and specific humidity derived from the North American Land Data Assimilation System Phase 2 (NLDAS-2), a health care access proxy defined as the distance from each ZIP code centroid to the nearest hospital identified from the American Hospital Association Annual Survey Database, and county-level mean body mass index and adult smoking prevalence derived from the Behavioral Risk Factor Surveillance System (BRFSS). Models also included calendar year and U.S. region (Northeast, Southeast, Midwest, Southwest, and West; Figure S1) fixed effects to account for temporal trends and spatial heterogeneity. All time-varying covariates were updated annually and entered as linear terms unless otherwise specified.

### Statistical analysis

For each outcome we fitted stratified Cox proportional-hazards models with years since cohort entry as the underlying time scale. Separate models were fitted for incident AD and incident ADRD using their corresponding cohorts. Baseline hazards were stratified by age at entry in 5-year groups, sex, race, and Medicaid eligibility.^11^ This approach was consistent with prior Medicare-based dementia analyses and allowed the baseline hazard to vary flexibly across strata without imposing proportional hazards assumptions for these variables. Robust standard errors clustered at the ZIP code level were used to account for residual within-ZIP correlation. Each model included both the smoke and corresponding non-smoke exposure terms simultaneously. Primary HRs and 95% CIs are reported per 1 µg/m^3^ and per IQR.

To evaluate potential nonlinearity in concentration-response relationships, we fitted penalized regression splines for smoke PM_2.5_ mass and smoke carbon separately for AD and ADRD. We assessed effect modification of the smoke associations by sex, race, age at cohort entry, Medicaid eligibility, geographic region, and baseline comorbidities, including stroke, hypertension, and depression. To evaluate the effect modification, we included multiplicative interaction terms between smoke exposure and each potential effect modifier in the fully adjusted model. Wald chi-square tests were used to assess statistical heterogeneity. All interaction terms were tested jointly for effect modifiers with more than two categories.

We also conducted sensitivity analyses to evaluate the robustness of the primary findings by: (1) using 1-, 2-, and 5-year moving-average exposure windows, (2) restricting the cohort to beneficiaries who did not change residential ZIP code during follow-up (approximately 86.5% of beneficiaries), (3) extending the outcome washout period from 3 to 5 years, (4) evaluating smoke OC, smoke EC, and smoke non-carbonaceous PM_2.5_ separately while adjusting for their corresponding non-smoke components, (5) applying stabilized inverse probability-of-censoring weights to account for censoring by death, as a way to account for censoring by death (S1.5), (6) replacing continuous smoke PM_2.5_ concentrations with annual counts of smoke days defined using thresholds of greater than 0, 1, and 2.5 µg/m^3^, (7) evaluating incident ADRD among beneficiaries without an AD-specific diagnosis (hereafter non-AD ADRD) as an alternative outcome, and (8) adjusting for claims-based indicators of tobacco use disorder, alcohol use disorder, and drug use disorder obtained from Medicare. Additional details and implementation of these sensitivity analyses are provided in Supplementary Section S1. All analyses were conducted in R version 4.1.2 on the Rollins High Performance Computing Cluster at Emory University.

### Attributable burden estimation

We translated the estimated HRs into the number of incident cases of AD and ADRD attributable to wildfire smoke. We limited this estimation to Medicare beneficiaries aged 65 to 100 years who resided in the CONUS and were continuously enrolled in fee-for-service Medicare with both Part A and Part B coverage, but without a required 3-year washout period. For each ZIP code and year, we combined primary effect estimates, 3-year moving average ZIP code-level exposure, and Medicare AD and ADRD incidence counts in a Monte Carlo simulation. Because separate component population-attributable fractions are not additive, we partitioned the total PM_2.5_-related burden into smoke and non-smoke PM_2.5_ and then allocated the smoke burden to PM_2.5_ carbon and the non-carbonaceous fraction using a Shapley attribution (details in Supplement Section S2).

## Results

### Study cohort and exposure distributions

A total of 39,678,630 Medicare beneficiaries met the inclusion criteria and contributed person-time to the cohorts for AD and ADRD (**Table 1**). From 2005 through 2019, the AD cohort accumulated 258.5 million person-years and 3,501,414 incident cases over a median follow-up of 6 years. The ADRD cohort accumulated 248.6 million person-years and 8,377,416 incident cases over a median follow-up of 5 years. At cohort entry, 69.4% of beneficiaries were aged <75 years, 54.5% were female, 85.9% were White, 7.6% were Black, and 13.9% were Medicaid dual-eligible. Hypertension was the most common comorbidity (64.1%), followed by depression (16.4%) and stroke (7.5%), while 32.3% had none of these three conditions. Geographically, the largest proportion of beneficiaries resided in the Southeast (31.1%).

**Table 1.** Baseline characteristics and exposure distributions of the AD and ADRD inception cohorts, U.S. Medicare beneficiaries, 2005 through 2019.

| Characteristic | AD cohort | ADRD cohort |
| --- | --- | --- |
| **Beneficiaries (shared denominator)** | 39,678,630 | 39,678,630 |
| **Incident events** | 3,501,414 | 8,377,416 |
| **Person-years (million)** | 258.5 | 248.6 |
| **Median follow-up (years)** | 6 | 5 |
| **Age at entry, number (%)** |  |  |
| <75 years | 27,542,701 (69.4) | 27,542,701 (69.4) |
| ≥75 years | 12,135,929 (30.6) | 12,135,929 (30.6) |
| **Sex, number (%)** |  |  |
| Male | 18,073,396 (45.5) | 18,073,396 (45.5) |
| Female | 21,605,234 (54.5) | 21,605,234 (54.5) |
| **Race, number (%)** |  |  |
| White | 34,098,628 (85.9) | 34,098,628 (85.9) |
| Black | 3,034,692 (7.6) | 3,034,692 (7.6) |
| Other<sup>1</sup> | 2,545,310 (6.4) | 2,545,310 (6.4) |
| **Medicaid dual-eligibility, number (%)** |  |  |
| Dual-eligible | 5,521,967 (13.9) | 5,521,967 (13.9) |
| Not dual-eligible | 34,156,663 (86.1) | 34,156,663 (86.1) |
| **Geographic region<sup>2</sup>, number (%)** |  |  |
| Northeast | 7,427,310 (18.7) | 7,427,310 (18.7) |
| Southeast | 12,342,467 (31.1) | 12,342,467 (31.1) |
| Midwest | 9,502,990 (23.9) | 9,502,990 (23.9) |
| Southwest | 4,222,246 (10.6) | 4,222,246 (10.6) |
| West | 6,183,617 (15.6) | 6,183,617 (15.6) |
| **Baseline comorbidity, number (%)** |  |  |
| Stroke | 2,977,783 (7.5) | 2,977,783 (7.5) |
| Hypertension | 25,419,427 (64.1) | 25,419,427 (64.1) |
| Depression | 6,491,062 (16.4) | 6,491,062 (16.4) |
| None of the three above | 12,807,542 (32.3) | 12,807,542 (32.3) |
| **3-year average exposure<sup>3</sup>, mean (SD), µg/m<sup>3</sup>** |  |  |
| Smoke PM<sub>2.5</sub> | 0.440 (0.387) | 0.439 (0.386) |
| Smoke carbon | 0.308 (0.214) | 0.307 (0.214) |
| Smoke OC | 0.256 (0.179) | 0.256 (0.178) |
| Smoke EC | 0.052 (0.040) | 0.051 (0.040) |
| Non-smoke PM<sub>2.5</sub> | 9.397 (2.229) | 9.417 (2.236) |
| Non-smoke carbon | 2.948 (0.830) | 2.952 (0.832) |

<sup>1</sup>Other indicates Asian, Hispanic, Native American, Pacific Islander, and multiracial individuals.
<sup>2</sup>Regions were obtained based on the first follow-up year.
<sup>3</sup>Exposure was population-weighted based on the LandScan global population database.
**Note:** The AD and ADRD inception cohorts were constructed from the same 39,678,630 eligible Medicare beneficiaries. The two cohorts differed in accumulated person-time, event counts, and median follow-up because of outcome-specific event definitions and censoring. Abbreviations: AD, Alzheimer's disease; ADRD, Alzheimer's disease and related dementias; EC, elemental carbon; IQR, interquartile range; OC, organic carbon; PM<sub>2.5</sub>, fine particulate matter with aerodynamic diameter of 2.5 µm or smaller; SD, standard deviation.

Exposure distributions were nearly identical between the AD and ADRD cohorts (**Table 1**). Across beneficiaries in the contiguous U.S., the mean 3-year smoke PM_2.5_ mass concentration was 0.440 µg/m^3^ (standard deviation [SD], 0.387), with an IQR of 0.322 µg/m^3^. The mean 3-year smoke carbon concentration was 0.308 µg/m^3^ (SD, 0.214), with an IQR of 0.260 µg/m^3^. Non-smoke concentrations were substantially higher in absolute terms, with mean concentrations of 9.40 µg/m^3^ for non-smoke PM_2.5_ mass and 2.95 µg/m^3^ for non-smoke carbon, and IQRs of 3.04 and 0.90 µg/m^3^, respectively. Smoke PM_2.5_ and smoke OC/EC were spatially concentrated in the western U.S., particularly in California, Oregon, Washington, Idaho, Montana, and the northern Rocky Mountain region (**Figure 1** and Figure S2). In contrast, non-smoke PM_2.5_, OC, and EC were highest across the eastern U.S. and in major urban corridors. Detailed exposure distributions are summarized in Tables S3 and S4.

**Figure 1.**
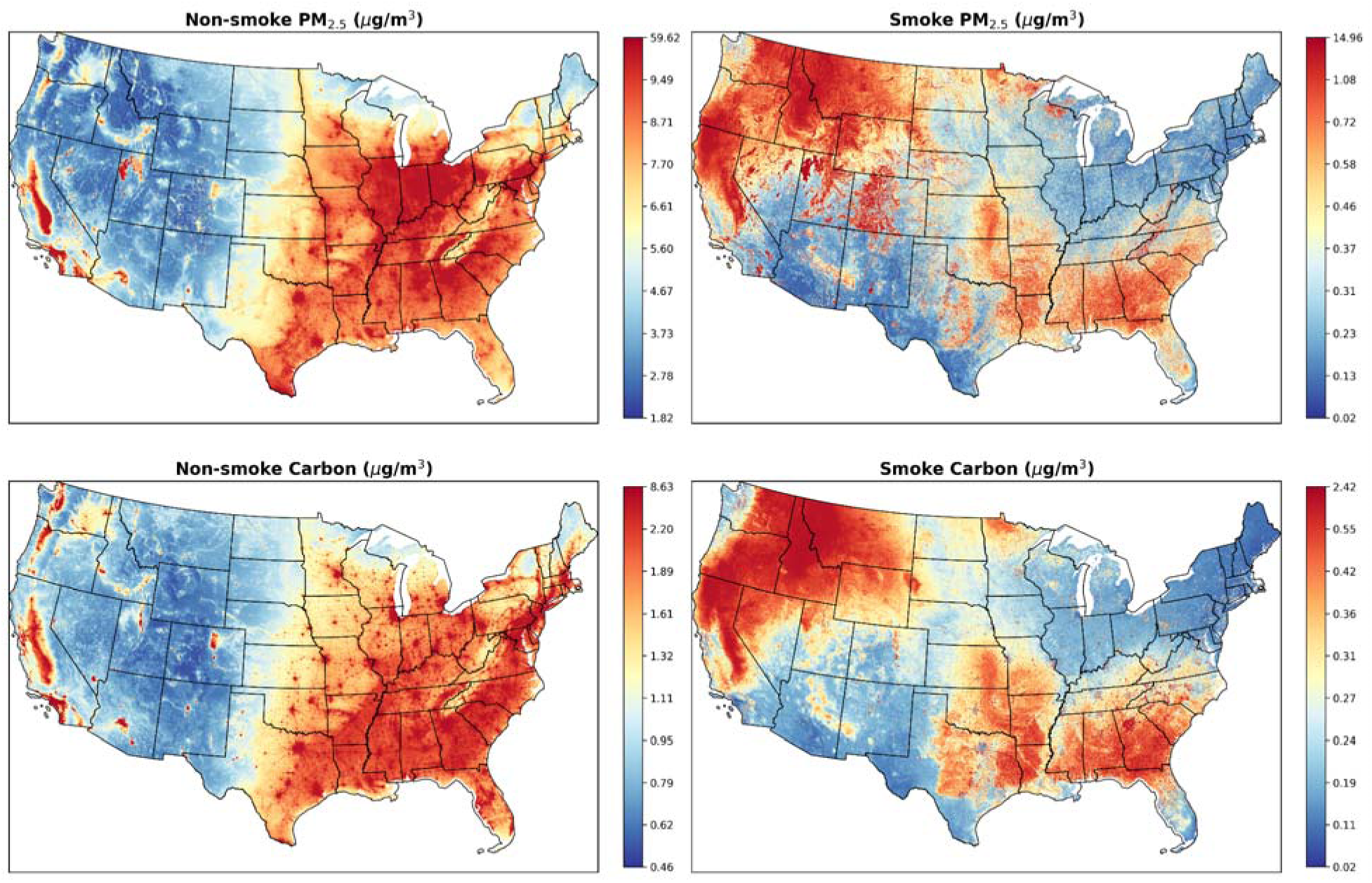
Mean annual concentrations (µg/m^3^) of non-smoke (left) and smoke (right) PM_2.5_ mass, and carbon (OC+EC) across the CONUS, 2002-2019. Mapped with 1-km spatial resolution.

### Main associations of long-term smoke exposure with incident AD and ADRD

In fully adjusted stratified Cox models, higher long-term exposure to smoke PM_2.5_ mass and smoke carbon was associated with higher incidence of both AD and ADRD (**Figure 2** and Table S5). For incident AD, the HR was 1.015 (95% CI: 1.005-1.026) per 1 µg/m^3^ increase in smoke PM_2.5_ and 1.063 (1.042-1.085) per 1 µg/m^3^ increase in smoke carbon. The corresponding HRs for non-smoke PM_2.5_ and non-smoke carbon were 1.028 (1.025-1.030) and 1.076 (1.069-1.082), respectively. For incident ADRD, the smoke-related associations were stronger than those observed for AD, with HRs of 1.029 (1.024-1.035) for smoke PM_2.5_ and 1.090 (1.077-1.103) for smoke carbon. The corresponding HRs for non-smoke PM_2.5_ and non-smoke carbon were 1.015 (1.014-1.017) and 1.034 (1.030-1.038), respectively. When scaled to their respective IQRs, associations with both smoke exposures remained positive. For AD, the per-IQR HRs were 1.005 (1.002-1.008) for smoke PM_2.5_ and 1.016 (1.011-1.021) for smoke carbon. For ADRD, the corresponding HRs were 1.009 (1.008-1.011) for smoke PM_2.5_ and 1.023 (1.019-1.026) for smoke carbon. Because smoke concentrations had narrower distributions, their per-IQR HRs were smaller than the corresponding estimates for non-smoke exposures for both outcomes (**Figure 2B**). The per-IQR HRs (Table S5) for non-smoke PM2.5 and non-smoke carbon were 1.086 (1.079-1.094) and 1.068 (1.062-1.073) for AD, and 1.048 (1.043-1.053) and 1.031 (1.027-1.034) for ADRD.

**Figure 2.**
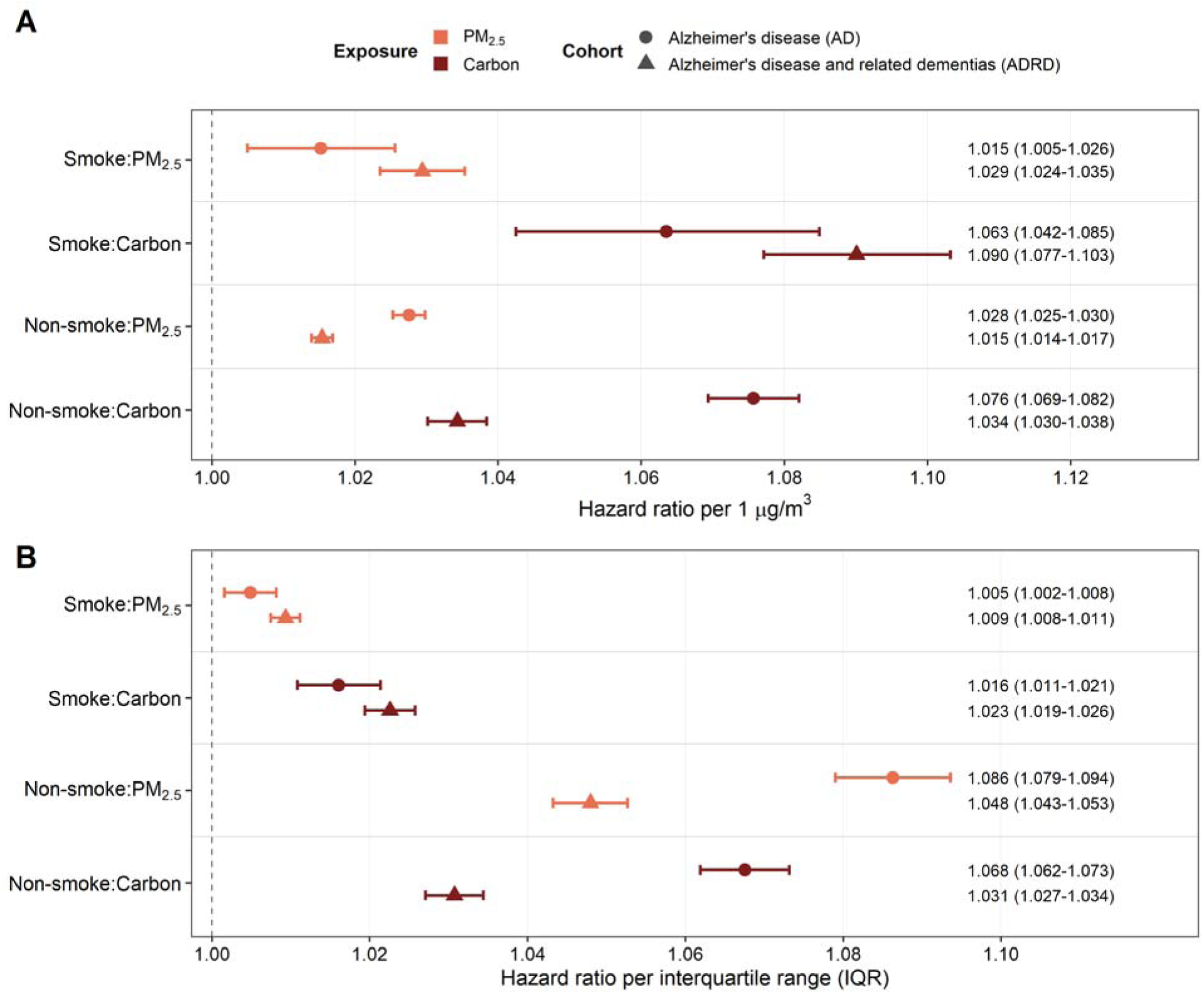
Associations of smoke and non-smoke PM_2.5_ and carbon exposures with incident AD and ADRD. (A) Per 1 µg/m^3^ increase. (B) Per IQR increase.

### Concentration-response relationships

Penalized spline models suggested nonlinear concentration-response relationships between long-term smoke exposure and incident AD and ADRD (**Figure 3**). For both outcomes, the estimated curves increased gradually at concentrations below approximately 0.50 µg/m^3^ for smoke PM_2.5_ and 0.25 µg/m^3^ for smoke carbon. Across intermediate exposure ranges, approximately 0.50 to 1.40 µg/m^3^ for smoke PM_2.5_ and 0.25 to 0.65 µg/m^3^ for smoke carbon, the slopes became steeper. At higher concentrations, the curves generally flattened or declined, with wider confidence intervals, consistent with fewer observations in the upper tail of the exposure distributions.

**Figure 3.**
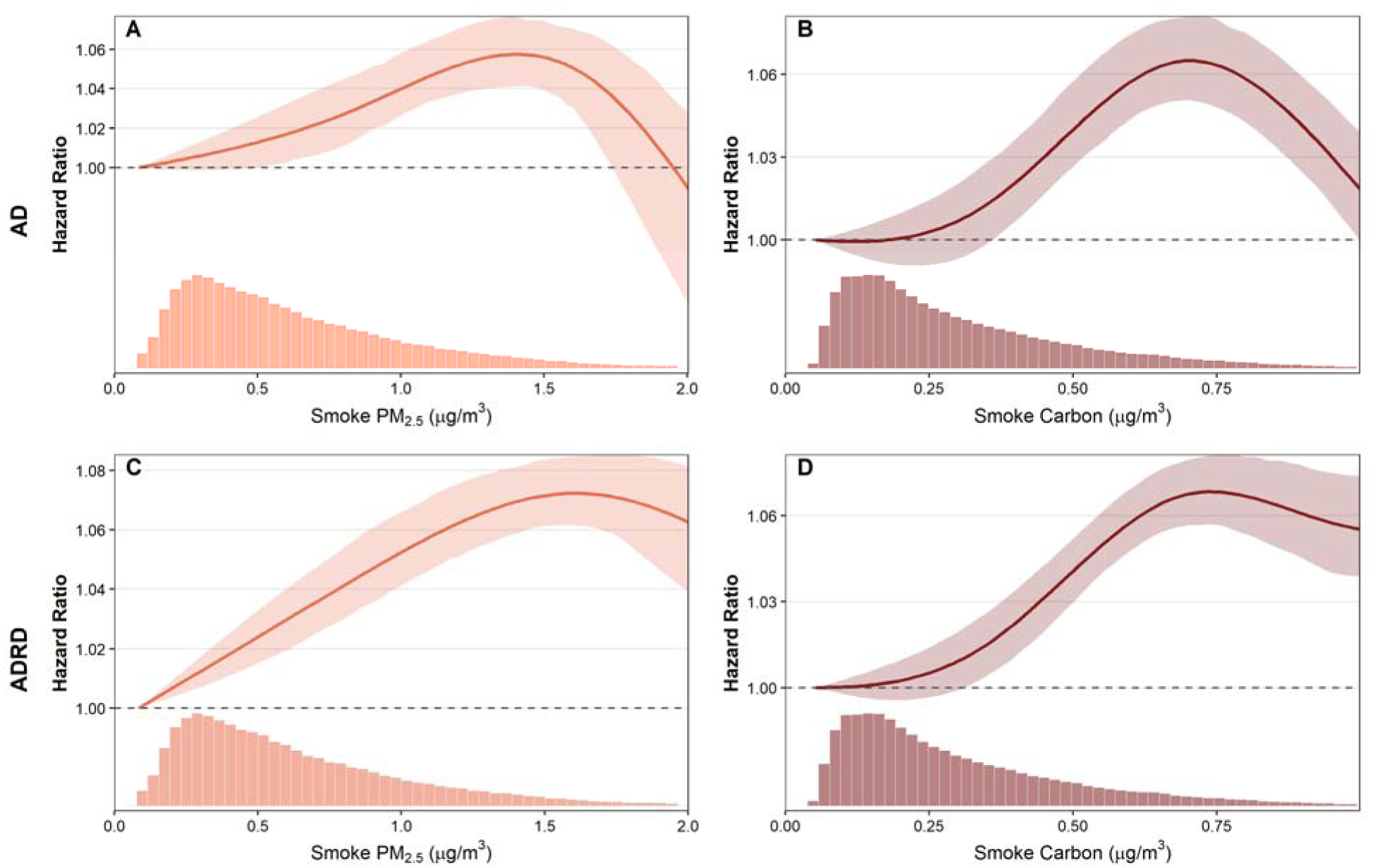
Concentration-response relationships between 3-year average wildfire smoke PM_2.5_ and smoke carbon and incident AD and incident ADRD. Note: Curves are displayed from the 1st to 99th percentile of each exposure distribution. Hazard ratios are referenced to the 1st percentile of each exposure. Shaded bands indicate 95% confidence intervals.

### Effect modification

Subgroup analyses suggested heterogeneity in the associations of smoke PM_2.5_ and smoke carbon with incident AD and ADRD (**Figure 4** and Table S6). The most consistent heterogeneity was observed by geographic region and baseline comorbidity status. Regional analyses showed the strongest positive associations in the Northeast for both outcomes, with smaller positive associations in the Southeast. Estimates in the Midwest and West were generally null or inverse. In analyses stratified by baseline clinical status, associations were generally stronger among beneficiaries without stroke, hypertension, or depression than among those with these conditions. Other subgroup patterns were less consistent. Differences by sex were small. Associations tended to be more positive among beneficiaries entering the cohort before age 75 years and among non-dual-eligible beneficiaries. Race-stratified estimates were positive among White beneficiaries for both outcomes. Full subgroup-specific HRs, 95% CIs, and interaction *P*-values are reported in Table S6.

**Figure 4.**
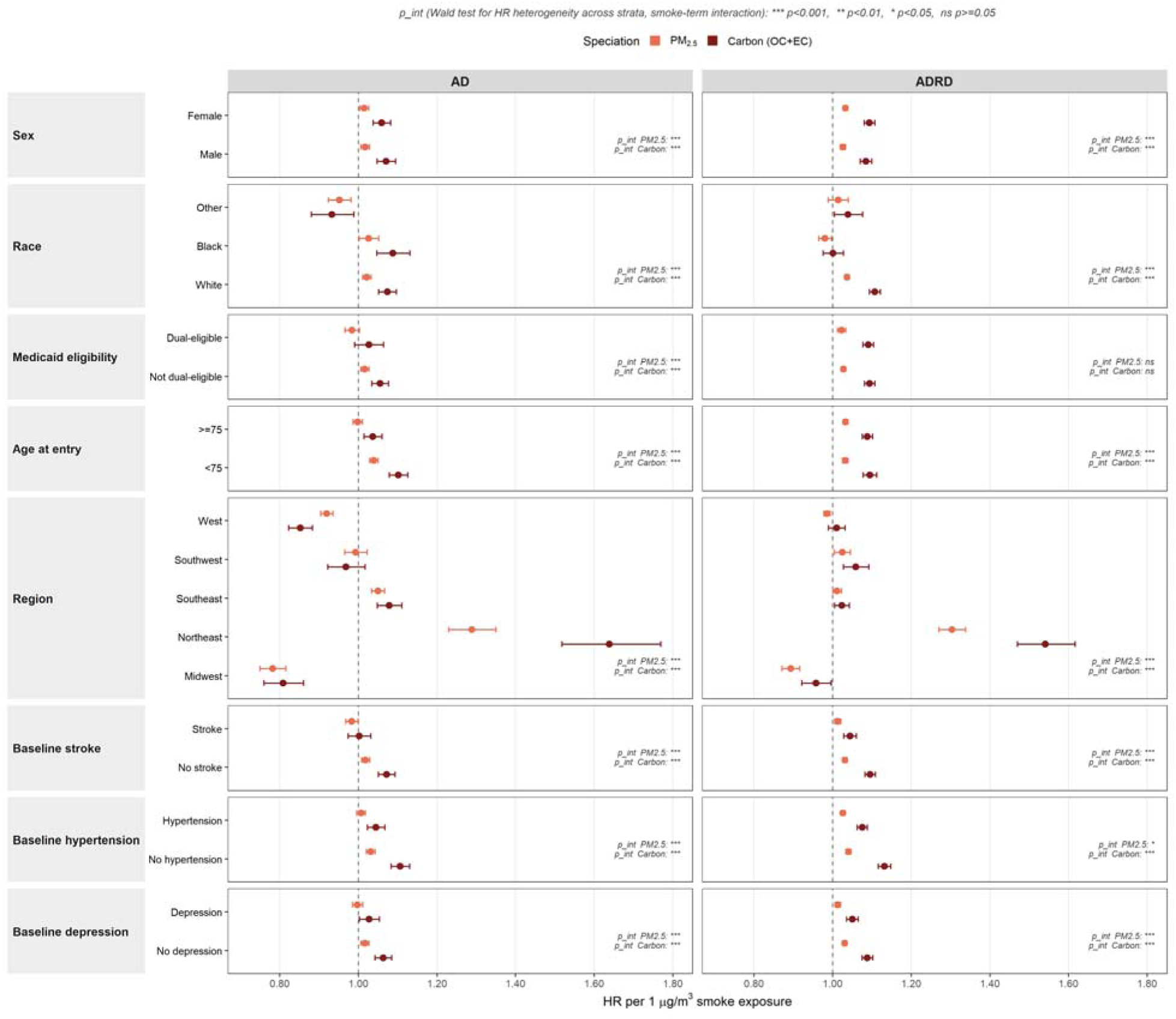
Effect modification of associations between wildfire smoke exposures and incident AD and ADRD.

### Sensitivity analyses

Primary associations were generally robust across the prespecified sensitivity analyses (Table S7). Associations with smoke PM_2.5_ and smoke carbon strengthened with longer cumulative exposure windows. Results showed stronger associations for the 5-year exposure window and weaker associations for the 1-year and 2-year exposure windows. Restricting the cohort to residential non-movers yielded modestly smaller but still positive estimates, whereas the estimates were essentially unchanged when extending the washout period to 5 years. Stabilized inverse probability-of-censoring weighting for death produced estimates that were nearly identical to the primary estimates. In component-resolved models, both smoke OC and smoke EC were positively associated with AD and ADRD, whereas the non-carbonaceous fraction of smoke PM_2.5_ showed weak or marginal associations with either outcome. Analyses using smoke-day counts showed positive associations across all three intensity thresholds, with stronger associations at higher smoke-day thresholds. In the analysis of non-AD ADRD, smoke PM_2.5_ and smoke carbon associations were stronger than those observed for AD and for ADRD. Associations were also generally consistent after additional adjustment for tobacco use disorder, alcohol use disorder, and drug use disorder.

### Attributable burden estimation

Long-term wildfire smoke PM_2.5_ was associated with a mean of 2,284 incident AD cases per year (95% CI: 733-3,833) and 11,142 incident ADRD cases per year (8,940-13,353) across the CONUS from 2005 through 2019, with smoke carbon accounting for most of this burden. These counts corresponded to 0.83% of incident AD cases and 1.69% of incident ADRD cases (Tables S8 and S9). In contrast, the mean annual numbers of cases attributable to non-smoke PM_2.5_ were higher, at 33,389 (30,995-35,988) for AD and 42,418 (38,779-46,638) for ADRD. Over the study period, the non-smoke PM_2.5_-attributable burden declined for both outcomes while the smoke-attributable burden rose for ADRD, so the smoke share of the PM_2.5_-attributable burden increased from 3.4% to 17.3% for AD and from 10.6% to 42.1% for ADRD (Figure S3). The smoke-attributable incidence rate was highest in the Mountain West and the Southeast and lowest in the Northeast, ranging across states from 3.6 to 19.5 attributable AD cases and from 19.3 to 120.1 attributable ADRD cases per 100,000 person-years (**Figure 5**).

**Figure 5.**
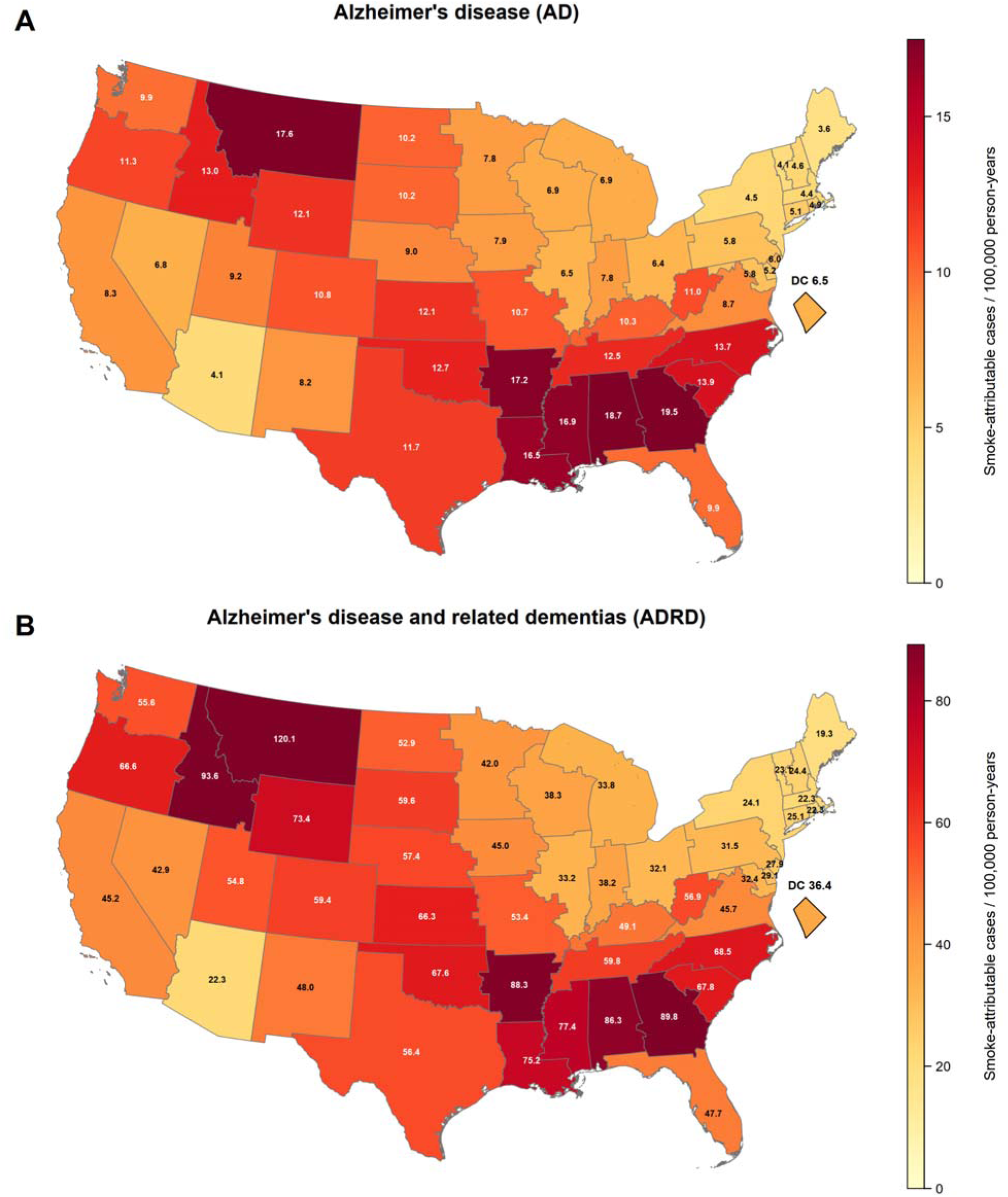
State-level annual incidence rates of AD and ADRD attributable to wildfire smoke PM_2.5_ mass, per 100,000 person-years.

## Discussion

In this nationwide cohort of older U.S. adults, long-term exposure to wildfire smoke PM_2.5_ was associated with higher incidence of both AD and ADRD. Smoke carbon was also positively associated with both outcomes and had larger per-unit-mass associations than smoke PM_2.5_. Within the broader ADRD endpoint, the per-unit-mass associations for smoke-related exposures were larger than those for their non-smoke counterparts. These comparisons should be interpreted cautiously because a 1 µg/m^3^ increase represented a very large and atypical contrast for smoke. The per-unit estimates provide standardized summary associations rather than effects that remain constant across the full exposure range. The IQR-based estimates and exposure-response curves therefore provide complementary information that more closely reflects the observed exposure distributions. To our knowledge, this study provides the first national, long-term, chemically resolved evaluation of wildfire smoke PM_2.5_ and smoke carbon in relation to incident AD and ADRD. The findings suggest that long-term wildfire smoke exposure, particularly its carbonaceous fraction, may contribute to neurodegenerative disease risk among older adults.

Prior epidemiological evidence on long-term wildfire-related particulate matter exposure and dementia remains limited and heterogeneous in design. In the U.S. Health and Retirement Study, Zhang et al. reported that source-apportioned wildfire PM_2.5_ at a resolution of approximately 50 × 70 km was associated with incident dementia after adjustment for PM_2.5_ from other sources and co-pollutants (HR per IQR of 0.6 µg/m^3^ in the 10-year mean, 1.05; 95% CI, 1.02-1.08; HR per 1 µg/m^3^, 1.09; 95% CI, 1.03-1.15).^15^ In northern Sweden, a smaller longitudinal cohort linked modeled PM_2.5_ from local residential wood burning (mean, 0.77 µg/m^3^; SD, 0.30) to incident dementia (HR per 1 µg/m^3^, 1.55; 95% CI, 1.00-2.41), providing evidence on chronic exposure to PM_2.5_ from residential wood combustion.^30^ More recently, Elser et al. evaluated 3-year average wildfire and non-wildfire PM_2.5_ in a large Kaiser Permanente Southern California cohort. The corrected primary estimate for wildfire PM_2.5_ was not statistically significant (odds ratio per 1 µg/m^3^, 1.12; 95% CI, 0.98-1.28), although subgroup associations were observed among participants younger than 75 years.^17^ Short-term smoke studies have also examined acute outcomes, including hospitalization among individuals with prevalent ADRD, but these endpoints address acute clinical exacerbation rather than incident neurodegenerative disease.^16^ Our study extends this literature by evaluating long-term smoke-specific exposure in a national Medicare cohort, applying mutual adjustment for smoke and non-smoke PM_2.5_, and examining smoke carbon as a major constituent of biomass-burning aerosol.

The positive associations observed for smoke PM_2.5_ and smoke carbon are biologically plausible given the chemical characteristics of biomass-combustion aerosol. Wildfire smoke PM_2.5_ is enriched in carbonaceous material, including OC from primary biomass-combustion emissions and secondary organic aerosol formation, as well as EC formed during incomplete combustion.^21,31,32^ These particles can contain redox-active organic compounds and combustion-derived material that may promote oxidative stress, systemic inflammation, endothelial dysfunction, and neuroinflammatory responses relevant to dementia pathogenesis.^6,8,33,34^ Atmospheric aging during smoke transport may further alter organic aerosol composition through oxidation and secondary organic aerosol formation, potentially altering its oxidative potential.^35,36^ By contrast, non-smoke PM_2.5_ represents a more heterogeneous mixture of secondary inorganic aerosols, carbonaceous particles from traffic and other combustion sources, industrial emissions, crustal material, and other sources.^37^ Although non-smoke particles can also exhibit substantial oxidative potential, their carbonaceous fraction may differ from wildfire smoke carbon in molecular composition, atmospheric processing, and redox activity.^38,39^ Moreover, our component-resolved sensitivity analyses provided complementary evidence regarding the composition of smoke PM_2.5_. The non-carbonaceous fraction of smoke PM_2.5_ showed weaker associations than those for smoke carbon, suggesting an important role of carbonaceous material in the association between long-term exposure to wildfire smoke PM_2.5_ and incident AD and ADRD.

In this study, we found that the per-unit associations with ADRD for smoke PM_2.5_ and smoke carbon exceeded those for their non-smoke counterparts, whereas the pattern was the opposite for AD. We explored this contrast in a sensitivity analysis that defined a non-AD ADRD endpoint. This endpoint does not isolate non-AD dementia. Many beneficiaries with an ADRD diagnosis but no AD-specific code probably have AD pathology, so the group is enriched for, rather than restricted to, non-AD etiologies. The non-AD ADRD endpoint showed the largest smoke-related HRs (Table S7), which were also larger than their non-smoke counterparts. One possible explanation is that wildfire smoke may be associated with pathological processes captured by the broader ADRD endpoint but not by AD-specific coding. This pattern is consistent with contributions from vascular, mixed, Lewy body, and other non-AD processes included within the broader ADRD definition. This interpretation is supported by recent evidence from a comparable national Medicare cohort in which long-term wildfire smoke exposure was associated with incident stroke.^40^ It is also compatible with clinicopathologic studies showing that late-life dementia commonly reflects coexisting Alzheimer, vascular, and other brain pathologies rather than AD alone.^41,42^ Claims-based ascertainment may also be relevant to the weaker AD-specific findings. AD-specific ascertainment in claims data is less sensitive and more vulnerable to subtype misclassification than broader dementia ascertainment. In prior validation work, Medicare claims showed sensitivity and specificity of 0.85 and 0.89 for dementia overall, compared with 0.64 and 0.95 for AD, and a CERAD-linked study identified 79% of clinically diagnosed AD cases using AD-specific claims versus 87% when broader dementia codes were used.^27,43^ More recent validation studies likewise suggest that claims-based algorithms perform reasonably for dementia overall, whereas AD-specific codes are substantially less sensitive.^28,44^ Such misclassification may attenuate AD-specific associations and contribute to the weaker contrast between smoke and non-smoke exposures observed for the AD endpoint.

We also observed marked spatial heterogeneity in smoke associations (**Figure 4**). Associations were strongest in the Northeast for both AD and ADRD, remained positive in the Southwest and Southeast, and were null or inverse in the Midwest and West. These differences likely reflect regional differences in smoke sources, atmospheric processing, exposure patterns, population responses, and diagnostic practices. The Northeast frequently experiences smoke transported over long distances from western U.S. and Canadian fires. During transport, atmospheric aging can alter the chemical composition and oxidative properties of carbonaceous particles, while the broad geographic coverage of transported plumes can expose large populations with limited awareness of wildfire smoke or insufficient adoption of protective measures.^45,46^ In the Southeast, frequent prescribed burning produces recurrent and spatially distributed smoke exposure throughout much of the year.^47^ Collectively, these regional patterns suggest that the health impact of long-term smoke exposure is shaped not only by concentration, but also by fire smoke source, fuel composition, transport history, and the frequency and duration of exposure. However, these factors were not directly identified in our epidemiological models. The regional findings should therefore be interpreted as descriptive evidence of geographic heterogeneity rather than as source-specific comparisons. Resolving these regional differences will require exposure metrics that characterize smoke itself, including transport time and plume age, fuel type and burn regime, and speciated organic aerosol markers.

Smoke associations also varied across strata defined by baseline clinical status, age, race, and socioeconomic status. Larger estimates were observed more frequently among beneficiaries without selected baseline comorbidities, those entering the cohort before age 75 years, White beneficiaries, and beneficiaries who were not eligible for Medicaid. This is consistent with a recent Medicare analysis of total PM_2.5_ and incident AD, in which hypertension, stroke, and depression mediated only 1.6% to 4.2% of the association and modified it modestly at most, indicating that PM_2.5_ acts on AD risk largely independently of these comorbidities.^48^ In addition, these patterns may partly reflect differences in the extent to which ambient smoke translates into personal exposure. Variation in indoor infiltration, time spent outdoors, and protective behaviors may cause the same increment in estimated ambient concentration to correspond to different increments in personal smoke exposure across subgroups.^45,49^ Younger beneficiaries and those without selected baseline comorbidities may be more mobile and spend more time outdoors, potentially increasing their contact with ambient smoke.^50,51^ In contrast, individuals with established comorbidities may spend less time outdoors or adopt behaviors that reduce smoke exposure during periods of poor air quality.^52^ Differences by Medicaid eligibility, which served as a proxy for socioeconomic status, may also reflect socioeconomic variation in residential exposure patterns.

Higher-income populations may be more likely to reside in portions of the wildland-urban interface where ambient smoke concentrations can be elevated.^53^ Differences in exposure distributions may influence subgroup estimates when concentration-response relationships are nonlinear. Together, these findings suggest that long-term smoke exposure may be influenced by activity patterns and residential characteristics, which should be interpreted with caution as these factors were unavailable in our study.

The attributable burden analysis indicated that wildfire smoke represents an increasing share of the long-term PM_2.5_-related dementia burden among older U.S. adults and is emerging as a substantial public health contributor to AD and ADRD. Although smoke contributes a smaller share of total PM_2.5_ mass than non-smoke sources, its higher toxicity and widespread plumes yield a substantial burden. By contrast, non-smoke PM_2.5_ has declined with national air-quality improvement from on-road controls, power generation, and other sectors, so that smoke accounts for a growing proportion of the PM_2.5_-related dementia burden over time. Integrating wildfire smoke into air-quality and climate-adaptation planning, and prioritizing mitigation and exposure reduction in the regions carrying the highest attributable rates will be important for protecting brain health in an aging population.

This study has several strengths. First, to our knowledge, it is the first large, nationwide cohort-based analysis of incident AD and ADRD associated with long-term exposure to smoke-specific PM_2.5_ mass and its carbonaceous fraction. Second, the cohort of approximately 40 million Medicare beneficiaries provided substantial statistical power and enabled systematic evaluation of heterogeneity across sociodemographic, regional, and clinical subgroups. Third, we used daily exposure estimates at 1-km spatial resolution that distinguished smoke from non-smoke PM_2.5_ and provided OC and EC concentrations. This approach improved exposure characterization relative to studies relying on proxy metrics such as smoke days or plume indicators. Fourth, we constructed parallel outcome-specific inception cohorts for AD and ADRD and applied a common analytic framework with mutually adjusted smoke and non-smoke exposures, facilitating comparison across the two related but distinct dementia outcomes. Finally, the consistency of positive associations across sensitivity analyses strengthens the robustness of the findings and supports their relevance for wildfire smoke mitigation and dementia prevention strategies.

Several limitations should be acknowledged. First, AD and ADRD were identified from Medicare claims rather than clinical assessment or neuropathological confirmation. Although claims-based algorithms perform reasonably well for dementia overall, AD-specific coding has lower sensitivity and is more vulnerable to subtype misclassification. The CCW ADRD definition also represents a heterogeneous outcome, and we could not separately evaluate vascular, Lewy body, frontotemporal, mixed, or other dementia subtypes. Second, detailed individual-level information on smoking history, physical activity, education, occupational history, and other behavioral or clinical risk factors was unavailable. In sensitivity analyses, the associations remained generally consistent after further adjustment for claims-based indicators of tobacco use disorder, alcohol use disorder, and drug use disorder. However, these indicators likely identify more severe or clinically documented disorders and may not capture the broader range of behavioral patterns. Although we accounted for multiple individual and area-level covariates and prior Medicare research suggests that PM_2.5_ associations are relatively robust to additional adjustment for individual risk factors, residual confounding cannot be excluded.^54^ Third, smoke and non-smoke exposures were model-derived and subject to prediction error, uncertainty in source separation, and potential regional differences in model performance. Annual ZIP code-level exposure assignment also did not capture indoor infiltration, filtration, or activity patterns. The consistency of results among non-movers and under alternative exposure definitions supports the stability of the findings but does not eliminate exposure misclassification. Fourth, we did not adjust for wildfire-related gaseous pollutants, including O_3_, NO_2_, and VOCs, because nationally complete, long-term, high-resolution, and source-resolved estimates were unavailable. These pollutants may co-occur with wildfire smoke and contribute independently or jointly to neurovascular and neuroinflammatory responses. Consequently, residual co-pollutant confounding cannot be excluded, and the smoke PM_2.5_ estimates may partly reflect exposure to the broader wildfire pollution mixture. Finally, the study population was limited to older fee-for-service Medicare beneficiaries and may not fully represent Medicare Advantage enrollees or younger adults.

## Acknowledgements

This study was supported by the National Institutes of Health (NIH, **R01ES034175**). Federal funds accounted for 100% of the total project costs (US$2,899,176). The content is solely the responsibility of the authors and does not necessarily represent the official views of the NIH. This air quality research used resources of the Oak Ridge Leadership Computing Facility at the Oak Ridge National Laboratory, which is supported by the Office of Science of the U.S. Department of Energy under Contract No. DE-AC05-00OR22725.

## Code availability

The exposure estimates and analysis code supporting the findings of this study are available from the corresponding author upon reasonable request.

## Data availability

Medicare data are not publicly available and are subject to CMS data-use agreements. Access may be obtained through CMS following its standard application procedures.

## Author contributions

ZJ and YL conceived and designed the study. ZJ developed the methods, performed the research, and wrote the initial draft of this manuscript. ZJ and KX accessed and verified the data. GAF, MQ, and JSF contributed the smoke PM_2.5_ modeling. Other authors contributed to the interpretation of the results and assisted in revising the manuscript. All authors had full access to all the data in the study and had final responsibility for the decision to submit for publication.

