## Supplemental Information, Tables, and Figures for "Carbonaceous Components Underlying Wildfire Smoke PM_2.5_-Associated Alzheimer’s Disease and Related Dementias in Older US Adults"

**Supplementary Information**

**S1. Sensitivity analyses**

We conducted a series of prespecified sensitivity analyses to evaluate the robustness of the primary findings to the exposure averaging window, residential mobility, outcome washout period, smoke-component specification, censoring by death, alternative representations of recurring smoke exposure, and outcome definition. Unless otherwise specified, all sensitivity analyses retained the cohort eligibility criteria, underlying time scale, model stratification, and covariate adjustment used in the primary analyses.

**S1.1 Alternative exposure averaging windows**

The primary analyses used annually updated 3-year moving-average concentrations of smoke PM_2.5_ and smoke carbon. To evaluate the cumulative effects of smoke and whether the findings were sensitive to the selected exposure window, we repeated the primary models using 1-year, 2-year, and 5-year moving-average concentrations for both exposures.

**S1.2 Restriction to beneficiaries who did not move**

Exposure was assigned according to annually updated residential ZIP codes rather than precise residential addresses, which could introduce exposure misclassification, particularly if changes in residence were not captured immediately or if prior exposure histories were incompletely represented after relocation. To evaluate the influence of residential mobility, we repeated the primary analyses among beneficiaries whose residential ZIP code remained unchanged throughout follow-up.

**S1.3 Extended outcome washout period**

The primary outcome-specific cohorts used a 3-year washout period to exclude beneficiaries with previously identified AD or ADRD. However, dementia may remain undiagnosed or may not appear in Medicare claims during its early clinical course. We therefore reconstructed the outcome-specific cohorts using a 5-year washout period.

**S1.4 Component-specific exposure models**

To examine whether the observed associations differed across smoke components, we conducted separate analyses of smoke OC, smoke EC, and smoke non-carbonaceous PM_2.5_. Smoke non-carbonaceous PM_2.5_ was calculated as smoke PM_2.5_ mass minus the sum of smoke OC and smoke EC. Each smoke component was included with its corresponding non-smoke component in the same model. All component exposures were evaluated using the same 3-year exposure window and model specification as the primary analysis.

**S1.5 Censoring by death**

Death precludes the subsequent diagnosis of AD or ADRD and may be associated with both wildfire smoke exposure and factors related to dementia risk. Consequently, censoring beneficiaries at death in the primary cause-specific Cox models could result in informative censoring if the probability of remaining alive differed according to exposure and covariate history. To evaluate the potential influence of this competing event, we refitted the primary stratified Cox proportional hazards models using stabilized inverse probability-of-censoring weights for censoring by death.^1^

The weights were estimated in three steps. First, we constructed a beneficiary-year dataset including observations for each year in which a beneficiary was alive and at risk for the outcome at the beginning of the interval. We fitted a pooled annual logistic regression model for the probability of remaining alive through year $t$, conditional on being alive at the beginning of that year. The denominator model included the 3-year moving-average smoke and non-smoke exposure terms used in the corresponding primary model, the full set of individual-level and ZIP code-level covariates included in the primary specification, and the model stratification variables, including 5-year age-at-entry group, sex, race, and Medicaid eligibility. Time-varying exposures and covariates were updated annually in accordance with the primary analysis.

We then fitted a numerator logistic regression model that included only the stratification variables. This model estimated the stabilizing probability of remaining alive within each combination of age-at-entry group, sex, race, and Medicaid eligibility. For each beneficiary-year, the stabilized weight increment was calculated as the predicted survival probability from the numerator model divided by the corresponding predicted survival probability from the denominator model. The unstabilized weight increment was the reciprocal of the predicted survival probability from the denominator model.

Second, the annual stabilized weight increments were accumulated multiplicatively over follow-up. The weight assigned to beneficiary $i$ in year $t$ was calculated as the cumulative product of the annual stabilized weight increments from cohort entry through year $t$. The resulting cumulative weight represented the inverse of the estimated probability of remaining uncensored by death through that year, conditional on the measured exposure and covariate history, with stabilization by the baseline stratification variables.

Third, to limit the influence of extreme cumulative weights and potential violations of the positivity assumption, cumulative weights were truncated at the 1st and 99th percentiles of their distributions within the model strata. After truncation, the weights were tightly centered around one, with a mean of 1.00 and an SD of 0.03. We repeated the weighted analyses without truncation as an implementation check. Removing truncation produced nearly identical point estimates with slightly wider confidence intervals.

The weighted Cox models retained the underlying time scale, baseline-hazard stratification, exposure specification, and covariate adjustment used in the corresponding primary models.

**S1.6 Annual frequency of smoke days**

Continuous moving-average concentrations summarize the average contribution of smoke PM_2.5_ but may be sensitive to the modeling uncertainty and may not fully represent repeated smoke episodes. To evaluate recurring smoke exposure under different intensity thresholds, we constructed annual counts of smoke days using the daily smoke PM_2.5_ estimates from the exposure model. For each ZIP code and calendar year, we calculated the number of days with daily smoke PM_2.5_ concentrations: greater than 0 µg/m^3^, greater than 1 µg/m^3^, or greater than 2.5 µg/m^3^. The threshold greater than 0 µg/m^3^ represented any modeled smoke contribution, whereas the higher thresholds represented progressively more intense smoke days. The resulting annual counts were population-weighted and assigned according to each beneficiary’s residential ZIP code and entered as annually updated time-varying exposures. Separate models were fitted for each threshold using the same cohort, non-smoke PM_2.5_, outcome definition, model stratification, and covariate adjustment as the primary analyses.

**S1.7 Alternative dementia definition**

The primary ADRD outcome included AD and other qualifying dementia diagnoses. We therefore defined an additional outcome within the primary ADRD cohort to evaluate incident non-AD ADRD and assess whether the broader ADRD association was primarily attributable to AD-specific diagnoses. The non-AD ADRD outcome was evaluated within the primary ADRD cohort using the same covariates and model specification as the main analysis. Beneficiaries who received an AD-specific diagnosis at any time during follow-up, including those whose AD diagnosis followed an earlier ADRD diagnosis and who were therefore classified as AD in the main analyses, were not counted as non-AD ADRD events; their follow-up was censored at the date of the ADRD diagnosis. All other incident ADRD diagnoses were counted as non-AD ADRD events. This analysis evaluated associations with ADRD pathways not initially identified as AD, which may include vascular, mixed, Lewy body, frontotemporal, and other qualifying dementia diagnoses. However, the analysis should not be interpreted as definitive etiologic classification of dementia subtype. Many beneficiaries with an ADRD diagnosis and no AD-specific code are likely to have AD pathology, so this endpoint is enriched for, rather than restricted to, non-AD etiologies.

**S1.8 Claims-based behavioral indicators**

Medicare data did not include detailed individual-level measures of smoking history, alcohol consumption, drug use, or other health-related behaviors. To assess potential residual confounding, we conducted sensitivity analyses that further adjusted the primary models for claims-based indicators of tobacco use disorder, alcohol use disorder, and drug use disorder from the CCW Other Chronic or Potentially Disabling Conditions segment. These indicators were not included in the primary analyses because their identification periods differed from those of the standard annual CCW covariates. Moreover, claims-based indicators likely represent more severe or clinically documented disorders and may not adequately capture the broader patterns of tobacco, alcohol, and drug use in the Medicare population.

**S2. Attributable burden estimation**

We quantified ZIP code–year AD and ADRD incident cases attributable to smoke and non-smoke PM_2.5_ using a coherent counterfactual and parametric Monte Carlo simulation (10,000 iterations). For each ZIP code and year, we (i) computed the population attributable fraction (PAF), defined as the proportion of cases that would be avoided if smoke and non-smoke PM_2.5_ were reduced from their observed levels to a low counterfactual level; (ii) combined the smoke and non-smoke PAFs into a total PM_2.5_ PAF; and (iii) used a Shapley (average attributable fraction) decomposition to allocate the total PM_2.5_ burden first between smoke and non-smoke PM_2.5,_ and then (iv) allocated within the smoke PM_2.5_, among carbonaceous and the non-carbonaceous fraction.

When multiple, correlated exposures contribute to the same cases (e.g., smoke and non-smoke PM_2.5_, or carbon, and non-carbonaceous smoke), simple single-exposure PAFs do not sum to the combined burden and can even exceed 100%. To avoid double-counting, we used a Shapley (average attributable fraction) decomposition, which is equivalent to the “average attributable fraction” proposed by Gefeller and Eide.^2,3^ The Shapley idea is to imagine all possible orders of eliminating the exposures (e.g., eliminate smoke first, then non-smoke versus eliminate non-smoke first, then smoke).^4^ For each order, we compute how many cases are prevented when we remove a given exposure at the point it is switched off, after accounting for any exposures already removed. The Shapley share for an exposure is the average of these marginal contributions across all orders. This method has three properties: it treats exposures symmetrically (no arbitrary hierarchy), the component shares sum exactly to the total PAF (no double-counting), and if an exposure has no effect, it receives zero share. In practical terms, the Shapley share for smoke PM_2.5_ can be interpreted as the expected fraction of cases that would be prevented by eliminating smoke PM_2.5_ if the order in which smoke and non-smoke PM_2.5_ are eliminated were chosen at random. Within the smoke PM_2.5_ share, we apply the same logic to decompose the smoke burden into contributions from smoke carbon, and non-carbonaceous smoke.

Let $D_{ct}$ denote observed incident AD or ADRD cases in ZIP code $z$, year $t$. For smoke PM_2.5_, $C_{zt}^{SMK}$, we used a counterfactual of no smoke contribution (0 µg/m^3^). For non-smoke PM_2.5_, we used a fixed counterfactual concentration of 5 µg/m^3^ from the World Health Organization annual air quality guideline for PM_2.5_.^5^

We compared each ZIP code–year’s non-smoke concentration to this counterfactual, using the excess above 5 µg/m^3^:

$$C_{zt}^{NS,+}=max\{C_{zt}^{NS}-5, 0\}$$

For each iteration $s$, we obtained the coefficients from the Cox models: $\beta_{smk}$ for smoke PM_2.5_, $\beta_{ns}$ for non-smoke PM_2.5_, and $\beta_{A}$ and $\beta_{B}$ for smoke carbon, and non-carbonaceous matter. For each ZIP code-year and iteration, we draw log-slopes from normal distributions implied by the 95% CI:

$$\beta_{smk}^{\left( s \right)}\sim N\left( \hat{\beta}_{smk},SE_{smk}^{2} \right)$$

$$\beta_{non-smoke}^{(s)}\sim N(\hat{\beta}_{non-smoke},SE_{non-smoke}^{2})$$

$\beta_{A}^{(s)},\beta_{B}^{(s)},\beta_{C}^{(s)}$ were drawn similarly. The smoke PAF is:

$$PAF_{smk,zt}^{(s)}=1-\exp\{-\beta_{smk}^{\left( s \right)}C_{zt}^{SMK}\}$$

and the non-smoke PAF (relative to the 5 µg/m^3^ counterfactual) is:

$$PAF_{ns,zt}^{(s)}=1-exp\{-\beta_{ns}^{\left( s \right)}C_{zt}^{NS,+}\}$$

Because $PAF_{smk}$ and $PAF_{ns}$ are not additively separable, we first combined smoke and non-smoke on the relative-risk (multiplicative) scale so that the total fraction is:

$$PAF_{tot,zt}^{(s)}=1-(1-PAF_{smk,zt}^{(s)})(1-PAF_{ns,zt}^{(s)}).$$

We then apportioned the total between smoke and non-smoke using the Shapley attribution based on the PAF function:

$$f\left( smk,ns \right)=1-exp\{-(smk+ns)\}$$

with doses $smk=\beta_{smk}^{\left( s \right)}C_{zt}^{SMK}$ and $ns=\beta_{ns}^{\left( s \right)}C_{zt}^{NS,+}$. For each pollutant, its Shapley share$\phi$is the average marginal contribution $f(S\cup\left\{ j \right\}-f\left( S \right))$ over all orders of removing $\{smk,ns\}$. The Shapley share of smoke is the average marginal contribution over the $2!=2$coalition orders:

$$\phi_{smk,zt}^{(s)}=\frac{1}{2}\left[ f\left( smk,0 \right)-f(0,0) \right]+\frac{1}{2}\left[ f\left( smk,ns \right)-f\left( 0,ns \right) \right]=\frac{1}{2}(1-e^{-smk})(1+e^{-ns})$$

And the non-smoke Shapley share is $\phi_{ns,zt}^{(s)}=PAF_{tot,zt}^{(s)}-\phi_{smk,zt}^{(s)}$. $\phi$ is the assigned share of the pollutant, not a standalone PAF for that pollutant. Cases attributable to smoke and non-smoke PM_2.5_ are:

$$AD_{smk,zt}^{\left( s \right)}=\phi_{smk,zt}^{\left( s \right)}D_{zt}$$

$$AD_{ns,zt}^{(s)}=\phi_{ns,zt}^{(s)}D_{zt}$$

Within the smoke PM_2.5_ portion, we further decomposed the burden into contributions from carbon, and the non-carbonaceous fraction using the Shapley attribution. For any subset $S\subseteq\{A,B$, define the component value function:

$$f_{\theta}\left( S \right)=1-\exp\left( -\sum_{j\in S} \theta_{j} \right)$$

where $\theta_{A}=\beta_{A}C_{zt}^{A},\theta_{B}=\beta_{B}C_{zt}^{B}$. For each smoke component, its provisional Shapley share$\tilde{\phi}$is the average marginal contribution over all $2!=2$ orders of removing $\{A,B\}$:

$$\tilde{\phi}_{A,zt}^{(s)}=\frac{1}{2}\left[ f\left( \left\{ \theta_{A},0,0 \right\} \right)-f\left( 0,0,0 \right) \right]+\frac{1}{2}\left[ f\left( \left\{ \theta_{A},\theta_{B} \right\} \right)-f\left( 0,\theta_{B} \right) \right]$$

$$\tilde{\phi}_{A,zt}^{(s)}=\frac{1}{2}\left( 1-e^{-\theta_{A}} \right)[1+e^{-\theta_{B}}]$$

with analogous expressions for $\tilde{\phi}_{B}$. Let the provisional total PAF implied by the component HRs:

$$\tilde{PAF}_{smk,zt}^{(s)}=1-e^{-(\theta_{A}+\theta_{B})}$$

We then scaled the provisional shares so that their sum equals the burden attributed to smoke PM_2.5_. The final Shapley shares were scaled as:

$$\phi_{A/B}^{(s)}=\tilde{\phi}_{A/B}^{(s)}\frac{\phi_{smk,zt}^{(s)}}{\tilde{PAF}_{smk,zt}^{(s)}}$$

By construction, $\phi_{A}^{s}+\phi_{B}^{s}=\phi_{smk,zt}^{(s)}$(no overlap). The incident cases attributable to each smoke component are $AD_{A,zt}^{(s)}=\phi_{A,zt}^{(s)}D_{zt}$, etc.

Uncertainty was propagated via Monte Carlo draws with 10,000 iterations. For each iteration we sampled log-slopes. We computed ZIP code–year PAFs, Shapley shares, and attributable cases, and then aggregated to states and nationally. We report the posterior mean and 95% confidence intervals (2.5th-97.5th percentiles across draws).

**Supplementary Table**

**Table S1.** ICD-9 and ICD-10 codes used to define AD and ADRD in the Chronic Conditions Warehouse algorithm.

| **Outcome** | **ICD-9^1^ codes** | **ICD-10^1^ codes** |
| --- | --- | --- |
| AD | 331.0 (any diagnosis on the claim) | G30.0, G30.1, G30.8, G30.9 (any diagnosis on the claim) |
| ADRD (all-cause dementia) | 331.0, 331.11, 331.19, 331.2, 331.7, 290.0, 290.10, 290.11, 290.12, 290.13, 290.20, 290.21, 290.3, 290.40, 290.41, 290.42, 290.43, 294.0, 294.10, 294.11, 294.20, 294.21, 294.8, 797 (any diagnosis on the claim) | F01.50, F01.51, F02.80, F02.81, F03.90, F03.91, F04, F05, F06.1, F06.8, G13.8, G30.0, G30.1, G30.8, G30.9, G31.01, G31.09, G31.1, G31.2, G94, R41.81, R54 (any diagnosis on the claim) |

^1^ICD-10 codes are effective since 10/2015; effective dates for ICD-9 codes vary, but are valid through 09/2015.

**Table S2.** Pearson correlation between concentrations of smoke and non-smoke PM_2.5_, OC, and EC at annual 1-km level. 2005 through 2019.

|  | Smoke  PM_2.5_ | Smoke  OC | Smoke  EC | Non-smoke  PM_2.5_ | Non-smoke  OC | Non-smoke  EC |
| --- | --- | --- | --- | --- | --- | --- |
| Smoke PM_2.5_ | / | 0.784 | 0.543 | -0.262 | -0.143 | -0.214 |
| Smoke OC | 0.784 | / | 0.751 | -0.301 | -0.193 | -0.291 |
| Smoke EC | 0.543 | 0.751 | / | -0.064 | 0.110 | -0.014 |
| Non-smoke PM_2.5_ | -0.262 | -0.301 | -0.064 | / | 0.779 | 0.747 |
| Non-smoke OC | -0.143 | -0.193 | 0.110 | 0.779 | / | 0.912 |
| Non-smoke EC | -0.214 | -0.291 | -0.014 | 0.747 | 0.912 | / |

**Table S3.** Distributions of 3-year moving average exposure levels for the AD cohort (unit: µg/m^3^).

| **3-year Average Exposure^1^** | **1%** | **5%** | **25%** | **50%** | **75%** | **95%** | **99%** |
| --- | --- | --- | --- | --- | --- | --- | --- |
| Smoke PM_2.5_ | 0.085 | 0.124 | 0.214 | 0.333 | 0.536 | 1.065 | 2.005 |
| Smoke PM_2.5_ Carbon | 0.053 | 0.081 | 0.150 | 0.246 | 0.410 | 0.738 | 1.000 |
| Smoke OC | 0.042 | 0.067 | 0.124 | 0.206 | 0.341 | 0.613 | 0.841 |
| Smoke EC | 0.005 | 0.010 | 0.023 | 0.040 | 0.069 | 0.132 | 0.186 |
| Non-smoke PM_2.5_ | 5.021 | 6.030 | 7.782 | 9.181 | 10.826 | 13.418 | 14.937 |
| Non-smoke PM_2.5_ Carbon | 1.316 | 1.816 | 2.443 | 2.874 | 3.339 | 4.231 | 5.892 |
| Non-smoke OC | 1.101 | 1.474 | 1.920 | 2.197 | 2.478 | 3.118 | 4.431 |
| Non-smoke EC | 0.208 | 0.314 | 0.498 | 0.662 | 0.862 | 1.241 | 1.574 |

^1^Exposures were population-weighted based on the LandScan global population database.

**Table S4.** Distributions of 3-year moving average exposure levels for the ADRD cohort (unit: µg/m^3^).

| **3-year Average Exposure^1^** | **1%** | **5%** | **25%** | **50%** | **75%** | **95%** | **99%** |
| --- | --- | --- | --- | --- | --- | --- | --- |
| Smoke PM_2.5_ | 0.085 | 0.123 | 0.213 | 0.332 | 0.535 | 1.064 | 2.003 |
| Smoke PM_2.5_ Carbon | 0.053 | 0.081 | 0.150 | 0.245 | 0.408 | 0.737 | 0.999 |
| Smoke OC | 0.042 | 0.067 | 0.124 | 0.205 | 0.340 | 0.612 | 0.840 |
| Smoke EC | 0.005 | 0.010 | 0.023 | 0.040 | 0.069 | 0.131 | 0.185 |
| Non-smoke PM_2.5_ | 5.022 | 6.033 | 7.793 | 9.203 | 10.853 | 13.445 | 14.946 |
| Non-smoke PM_2.5_ Carbon | 1.315 | 1.816 | 2.446 | 2.879 | 3.344 | 4.237 | 5.903 |
| Non-smoke OC | 1.100 | 1.475 | 1.921 | 2.200 | 2.481 | 3.121 | 4.437 |
| Non-smoke EC | 0.207 | 0.314 | 0.499 | 0.664 | 0.864 | 1.245 | 1.578 |

^1^Exposures were population-weighted based on the LandScan global population database.

**Table S5.** Associations of 3-year average smoke and non-smoke PM_2.5_ and carbon with incident AD and ADRD among U.S. Medicare beneficiaries, 2005 through 2019.

| **Outcome** | **Exposure** | **HR per 1 µg/m^3^ (95% CI)** | **HR per IQR (95% CI)** | **IQR (µg/m^3^)** |
| --- | --- | --- | --- | --- |
| AD | Smoke PM_2.5_ | 1.015 (1.005-1.026) | 1.005 (1.002-1.008) | 0.322 |
|  | Non-smoke PM_2.5_ | 1.028 (1.025-1.030) | 1.086 (1.079-1.094) | 3.044 |
|  | Smoke carbon | 1.063 (1.042-1.085) | 1.016 (1.011-1.021) | 0.260 |
|  | Non-smoke carbon | 1.076 (1.069-1.082) | 1.068 (1.062-1.073) | 0.896 |
| ADRD | Smoke PM_2.5_ | 1.029 (1.024-1.035) | 1.009 (1.008-1.011) | 0.321 |
|  | Non-smoke PM_2.5_ | 1.015 (1.014-1.017) | 1.048 (1.043-1.053) | 3.060 |
|  | Smoke carbon | 1.090 (1.077-1.103) | 1.023 (1.019-1.026) | 0.259 |
|  | Non-smoke carbon | 1.034 (1.030-1.038) | 1.031 (1.028-1.035) | 0.898 |

**Table S6.** Stratum-specific hazard ratios and interaction *P*-values for effect modification of the smoke PM_2.5_ and smoke carbon associations with incident AD and incident ADRD.

| **Subgroup** | **AD, smoke PM_2.5_** | **AD, smoke carbon** | **ADRD, smoke PM_2.5_** | **ADRD, smoke carbon** |
| --- | --- | --- | --- | --- |
| **Sex** |  |  |  |  |
| Male | 1.017 (1.006-1.028) | 1.071 (1.047-1.094) | 1.026 (1.019-1.033) | 1.085 (1.070-1.100) |
| Female | 1.014 (1.003-1.026) | 1.059 (1.037-1.082) | 1.032 (1.026-1.038) | 1.094 (1.080-1.107) |
| *P-interaction* | <0.001 | <0.001 | <0.001 | <0.001 |
| **Race** |  |  |  |  |
| White | 1.021 (1.011-1.032) | 1.073 (1.051-1.096) | 1.036 (1.030-1.043) | 1.107 (1.093-1.121) |
| Black | 1.026 (1.001-1.051) | 1.088 (1.046-1.131) | 0.980 (0.964-0.996) | 1.001 (0.975-1.027) |
| Other | 0.952 (0.923-0.981) | 0.933 (0.880-0.988) | 1.014 (0.988-1.040) | 1.039 (1.004-1.076) |
| *P-interaction* | <0.001 | <0.001 | <0.001 | <0.001 |
| **Medicaid eligibility** | |  |  |  |
| Not dual-eligible | 1.016 (1.006-1.027) | 1.055 (1.034-1.076) | 1.027 (1.021-1.033) | 1.094 (1.081-1.107) |
| Dual-eligible | 0.984 (0.966-1.002) | 1.026 (0.990-1.064) | 1.022 (1.012-1.033) | 1.091 (1.077-1.104) |
| *P-interaction* | <0.001 | <0.001 | 0.092 | 0.561 |
| **Age at entry** |  |  |  |  |
| <75 years | 1.039 (1.029-1.050) | 1.102 (1.078-1.125) | 1.032 (1.025-1.039) | 1.094 (1.078-1.112) |
| ≥75 years | 0.998 (0.986-1.010) | 1.037 (1.014-1.060) | 1.033 (1.026-1.039) | 1.088 (1.075-1.101) |
| *P-interaction* | <0.001 | <0.001 | <0.001 | <0.001 |
| **Geographic region** | |  |  |  |
| Northeast | 1.288 (1.230-1.349) | 1.638 (1.518-1.769) | 1.304 (1.270-1.338) | 1.541 (1.470-1.616) |
| Southeast | 1.050 (1.033-1.066) | 1.078 (1.048-1.110) | 1.011 (1.000-1.022) | 1.023 (1.005-1.042) |
| Midwest | 0.782 (0.750-0.815) | 0.808 (0.759-0.860) | 0.893 (0.871-0.916) | 0.958 (0.921-0.996) |
| Southwest | 0.993 (0.965-1.022) | 0.968 (0.922-1.017) | 1.024 (1.004-1.045) | 1.059 (1.027-1.092) |
| West | 0.919 (0.904-0.935) | 0.852 (0.823-0.883) | 0.985 (0.978-0.993) | 1.010 (0.989-1.031) |
| *P-interaction* | <0.001 | <0.001 | <0.001 | <0.001 |
| **Baseline stroke** | |  |  |  |
| No stroke | 1.018 (1.008-1.028) | 1.071 (1.050-1.093) | 1.031 (1.025-1.037) | 1.095 (1.082-1.109) |
| Stroke | 0.983 (0.968-0.999) | 1.002 (0.974-1.032) | 1.013 (1.004-1.021) | 1.044 (1.028-1.060) |
| *P-interaction* | <0.001 | <0.001 | <0.001 | <0.001 |
| **Baseline hypertension** | |  |  |  |
| No hypertension | 1.031 (1.020-1.042) | 1.106 (1.083-1.130) | 1.040 (1.033-1.047) | 1.132 (1.116-1.148) |
| Hypertension | 1.007 (0.995-1.018) | 1.045 (1.023-1.067) | 1.026 (1.020-1.032) | 1.075 (1.062-1.088) |
| *P-interaction* | <0.001 | <0.001 | 0.015 | <0.001 |
| **Baseline depression** | |  |  |  |
| No depression | 1.016 (1.006-1.027) | 1.063 (1.042-1.085) | 1.030 (1.024-1.036) | 1.088 (1.075-1.102) |
| Depression | 0.998 (0.985-1.011) | 1.027 (1.002-1.053) | 1.013 (1.006-1.020) | 1.050 (1.035-1.065) |
| *P-interaction* | <0.001 | <0.001 | <0.001 | <0.001 |

**Table S7.** Sensitivity analyses results.

| **Sensitivity analysis** | **Exposure** | **AD** | **ADRD** |
| --- | --- | --- | --- |
| 1. Cumulative exposure window — per 1 µg/m^3^ | | |  |
| 1-year moving average | Smoke PM_2.5_ | 1.004 (0.997-1.011) | 1.012 (1.008-1.015) |
| 2-year moving average | Smoke PM_2.5_ | 1.011 (1.002-1.020) | 1.021 (1.016-1.026) |
| **3-year moving average (primary)** | **Smoke PM_2.5_** | **1.015 (1.005-1.026)** | **1.029 (1.024-1.035)** |
| 5-year moving average | Smoke PM_2.5_ | 1.017 (1.005-1.028) | 1.037 (1.030-1.043) |
| 1-year moving average | Smoke carbon | 1.054 (1.037-1.072) | 1.068 (1.057-1.078) |
| 2-year moving average | Smoke carbon | 1.062 (1.042-1.082) | 1.082 (1.070-1.094) |
| **3-year moving average (primary)** | **Smoke carbon** | **1.063 (1.042-1.085)** | **1.090 (1.077-1.103)** |
| 5-year moving average | Smoke carbon | 1.066 (1.044-1.089) | 1.095 (1.081-1.109) |
| 2. Cohort restriction — per 1 µg/m^3^ | | |  |
| Non-mover restriction | Smoke PM_2.5_ | 1.011 (1.002-1.020) | 1.024 (1.018-1.031) |
| Non-mover restriction | Smoke carbon | 1.045 (1.024-1.066) | 1.074 (1.061-1.087) |
| 3. Extended washout period — per 1 µg/m^3^ | | |  |
| 5-year washout | Smoke PM_2.5_ | 1.016 (1.006-1.026) | 1.029 (1.024-1.035) |
| 5-year washout | Smoke carbon | 1.063 (1.042-1.085) | 1.092 (1.079-1.105) |
| 4. Competing risk — per 1 µg/m^3^ | | |  |
| Censoring-by-death weighted | Smoke PM_2.5_ | 1.015 (1.005-1.026) | 1.029 (1.024-1.035) |
| Censoring-by-death weighted | Smoke carbon | 1.064 (1.043-1.085) | 1.090 (1.077-1.103) |
| 5. Component-resolved exposure — per 1 µg/m^3^ | | | |
| Component decomposition | Smoke OC | 1.053 (1.028-1.078) | 1.101 (1.086-1.117) |
| Component decomposition | Smoke EC | 1.731 (1.565-1.915) | 1.620 (1.522-1.724) |
| Component decomposition | Smoke non-carbon | 1.011 (1.002-1.020) | 1.005 (0.999-1.010) |
| 6. Smoke-day intensity — per 10 additional smoke days per year | | | |
| Class 1 (> 0 µg/m^3^) | Smoke days/year | 1.004 (1.003-1.005) | 1.004 (1.004-1.005) |
| Class 2 (> 1 µg/m^3^) | Smoke days/year | 1.006 (1.004-1.007) | 1.006 (1.005-1.007) |
| Class 3 (> 2.5 µg/m^3^) | Smoke days/year | 1.006 (1.004-1.008) | 1.007 (1.006-1.008) |
| 7. Non-AD ADRD sensitivity outcome — per 1 µg/m^3^ | | | |
| Non-AD ADRD outcome | Smoke PM_2.5_ | 1.031 (1.023-1.039) | |
| Non-AD ADRD outcome | Non-smoke PM_2.5_ | 1.005 (1.003-1.008) | |
| Non-AD ADRD outcome | Smoke carbon | 1.096 (1.080-1.112) | |
| Non-AD ADRD outcome | Non-smoke carbon | 1.010 (1.005-1.015) | |
| 8. Additional adjustment for claims-based tobacco, alcohol, and drug use disorders — per 1 µg/m^3^ | | | |
| Additional adjustment | Smoke PM_2.5_ | 1.016 (1.006-1.026) | 1.029 (1.024-1.035) |
| Additional adjustment | Smoke carbon | 1.061 (1.040-1.083) | 1.084 (1.071-1.097) |

**Table S8.** Annual attributable incident AD cases for non-smoke and smoke PM_2.5_ among Medicare older adults (2005-2019), with allocation to carbonaceous fraction.

| **Year** | **Total** | **Non-smoke PM_2.5_** | **Smoke PM_2.5_** | **Smoke Carbon** |
| --- | --- | --- | --- | --- |
| 2005 | 409,906 | 63,705 (59,137-68,486) | 2,219 (711-3,731) | 1,831 (578-3,177) |
| 2006 | 387,061 | 59,984 (55,697-64,506) | 2,303 (738-3,872) | 1,921 (607-3,322) |
| 2007 | 368,477 | 55,027 (51,104-59,219) | 2,516 (807-4,227) | 2,074 (655-3,594) |
| 2008 | 356,114 | 52,609 (48,904-56,675) | 3,049 (978-5,119) | 2,473 (781-4,300) |
| 2009 | 341,117 | 46,333 (43,061-49,963) | 3,212 (1,032-5,391) | 2,577 (813-4,493) |
| 2010 | 315,702 | 39,319 (36,520-42,419) | 2,948 (947-4,946) | 2,358 (743-4,115) |
| 2011 | 295,920 | 33,926 (31,478-36,598) | 2,455 (788-4,120) | 1,999 (632-3,475) |
| 2012 | 283,214 | 31,040 (28,800-33,500) | 2,491 (800-4,180) | 2,065 (654-3,572) |
| 2013 | 257,006 | 27,027 (25,075-29,178) | 2,361 (758-3,960) | 1,985 (630-3,421) |
| 2014 | 234,441 | 22,611 (20,971-24,422) | 2,246 (722-3,766) | 1,884 (598-3,248) |
| 2015 | 237,789 | 20,840 (19,310-22,505) | 2,050 (659-3,438) | 1,727 (548-2,976) |
| 2016 | 219,885 | 18,171 (16,832-19,626) | 1,846 (593-3,095) | 1,556 (494-2,681) |
| 2017 | 186,550 | 13,929 (12,900-15,051) | 1,612 (518-2,703) | 1,372 (435-2,358) |
| 2018 | 149,330 | 9,904 (9,179-10,718) | 1,621 (521-2,714) | 1,370 (435-2,357) |
| 2019 | 104,968 | 6,415 (5,951-6,950) | 1,337 (431-2,238) | 1,122 (356-1,933) |
| Average | 276,499 | 33,389 (30,995-35,988) | 2,284 (733-3,833) | 1,888 (597-3,268) |

**Table S9.** Annual attributable incident ADRD cases for non-smoke and smoke PM_2.5_ among Medicare older adults (2005-2019), with allocation to carbonaceous fraction.

| **Year** | **Total** | **Non-smoke PM_2.5_** | **Smoke PM_2.5_** | **Smoke Carbon** |
| --- | --- | --- | --- | --- |
| 2005 | 765,008 | 69,037 (63,017-75,621) | 8,218 (6,588-9,860) | 7,686 (6,081-9,394) |
| 2006 | 725,210 | 65,165 (59,509-71,416) | 8,512 (6,825-10,212) | 8,001 (6,346-9,751) |
| 2007 | 693,616 | 59,895 (54,735-65,712) | 9,325 (7,479-11,185) | 8,726 (6,910-10,651) |
| 2008 | 681,368 | 58,024 (53,122-63,780) | 11,503 (9,230-13,791) | 10,702 (8,458-13,088) |
| 2009 | 661,212 | 51,546 (47,210-56,729) | 12,296 (9,868-14,736) | 11,383 (8,977-13,956) |
| 2010 | 637,396 | 45,459 (41,614-50,041) | 11,725 (9,409-14,053) | 10,837 (8,539-13,300) |
| 2011 | 632,329 | 41,413 (37,859-45,555) | 10,329 (8,286-12,383) | 9,607 (7,589-11,764) |
| 2012 | 643,119 | 40,262 (36,818-44,316) | 11,083 (8,892-13,286) | 10,387 (8,234-12,666) |
| 2013 | 610,103 | 36,564 (33,440-40,261) | 11,007 (8,832-13,192) | 10,374 (8,243-12,616) |
| 2014 | 581,959 | 31,917 (29,185-35,158) | 10,963 (8,798-13,136) | 10,320 (8,196-12,556) |
| 2015 | 631,784 | 31,520 (28,785-34,693) | 10,711 (8,594-12,836) | 10,098 (8,021-12,281) |
| 2016 | 692,104 | 32,499 (29,668-35,770) | 11,383 (9,132-13,640) | 10,735 (8,528-13,055) |
| 2017 | 662,522 | 27,966 (25,531-30,795) | 11,163 (8,956-13,376) | 10,561 (8,400-12,827) |
| 2018 | 632,594 | 23,665 (21,645-26,121) | 13,426 (10,777-16,076) | 12,668 (10,068-15,397) |
| 2019 | 619,630 | 21,332 (19,548-23,596) | 15,489 (12,441-18,534) | 14,571 (11,570-17,727) |
| Average | 657,997 | 42,418 (38,779-46,638) | 11,142 (8,940-13,353) | 10,444 (8,277-12,735) |

Supplementary Figures

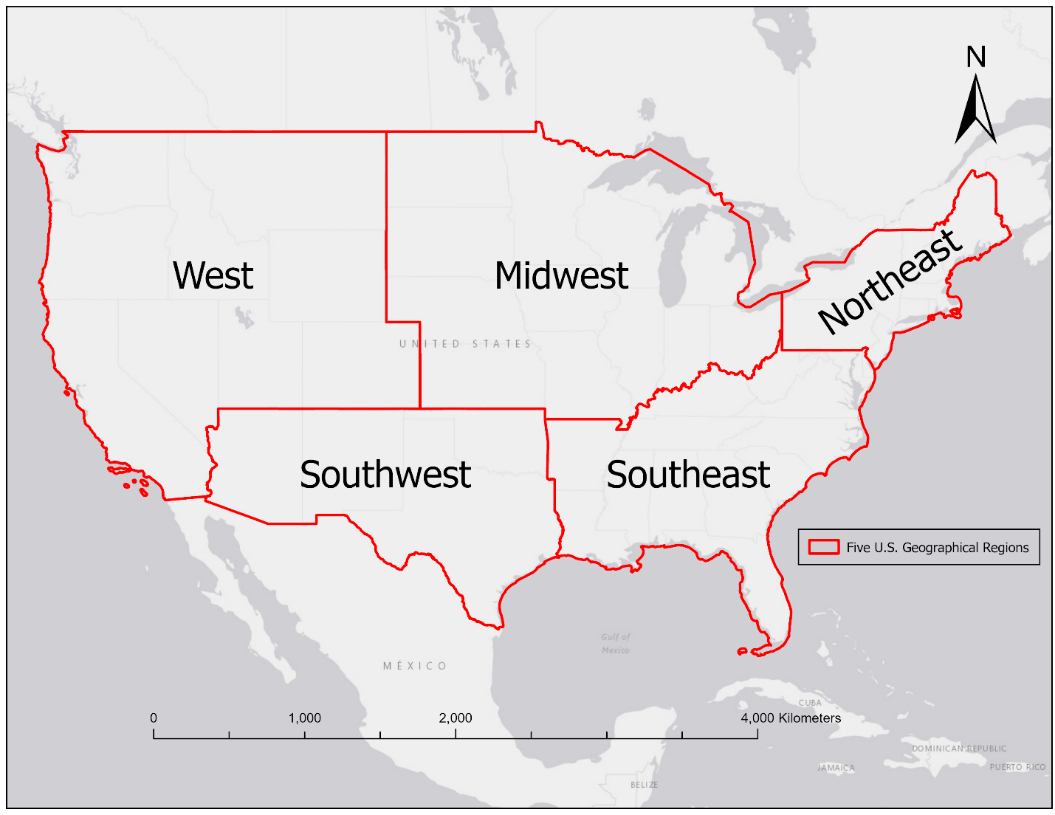

**Figure S1.** Five U.S. geographical regions used in this study (Northeast, Southeast, Midwest, West, Southwest).

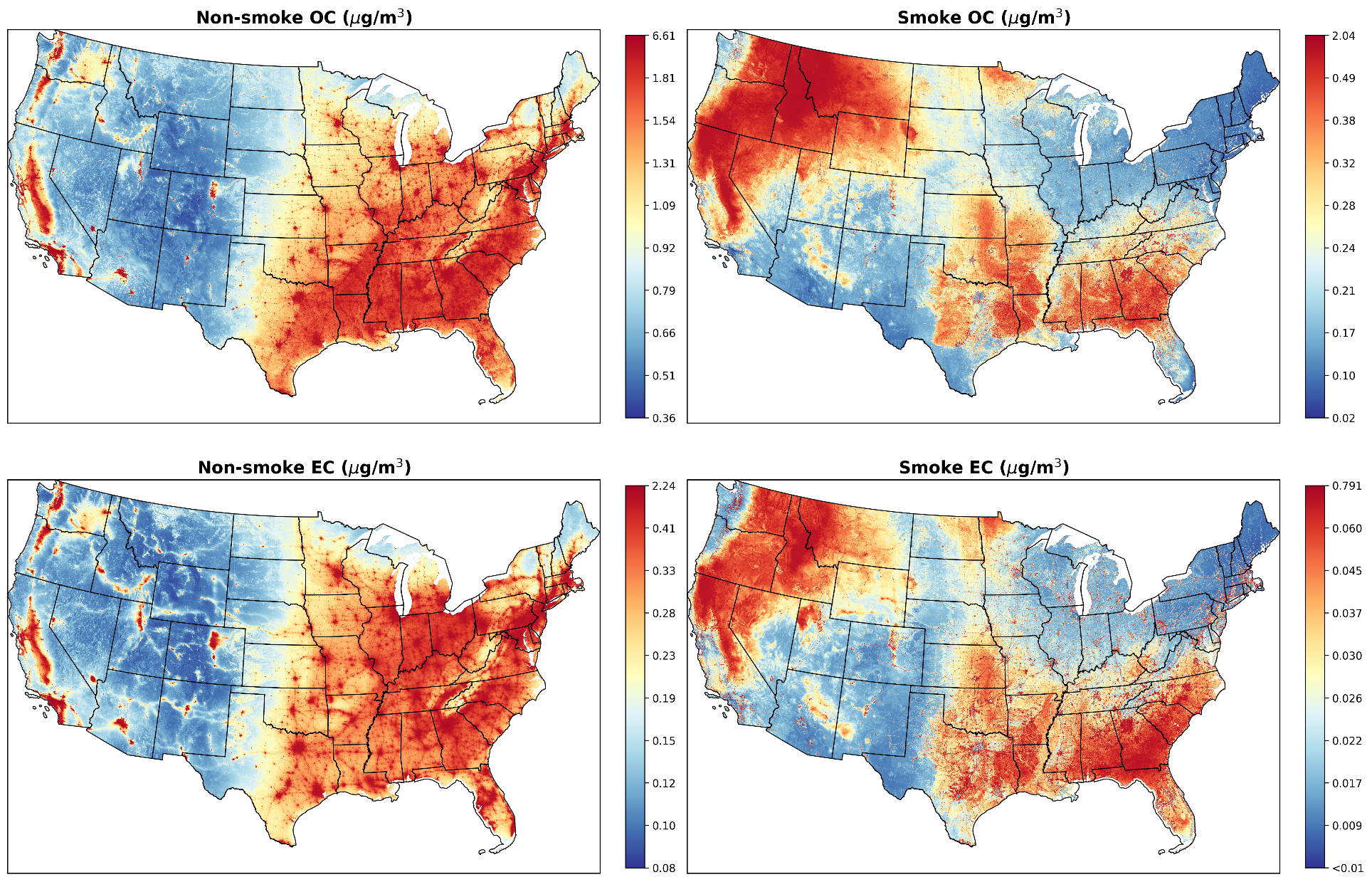

**Figure S2.** Mean annual concentrations (µg/m^3^) of non-smoke (left) and smoke (right) OC and EC across the CONUS, 2002-2019. Mapped with 1-km spatial resolution.

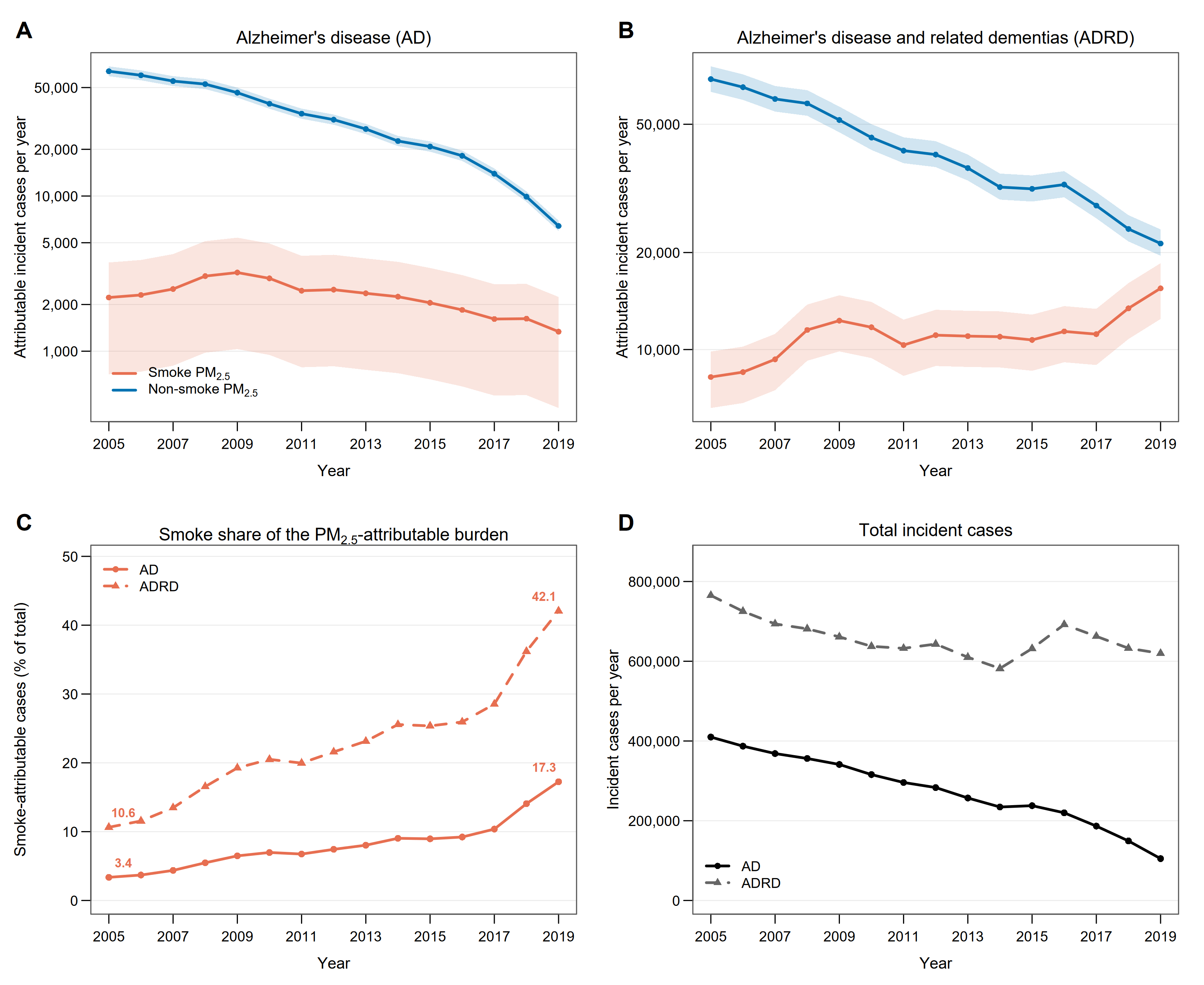

**Figure S3.** Annual incident AD and ADRD cases attributable to smoke and non-smoke PM_2.5_ among U.S. Medicare beneficiaries, 2005 through 2019. (A, B) Attributable cases per year with 95% CI, on a logarithmic axis, on which the intervals appear asymmetric although they are symmetric on the natural scale. (C) Smoke as a percentage of the PM_2.5_-attributable burden, with 2005 and 2019 values labeled. (D) Total incident cases per year. Estimation is described in Methods, and annual values are given in Supplementary Tables S8 and S9.

5	World Health Organization. *WHO global air quality guidelines: particulate matter (PM2.5 and PM10), ozone, nitrogen dioxide, sulfur dioxide and carbon monoxide: executive summary*. (World Health Organization, 2021).
